# Re-classification and Weighting of Multiple Causes of Death: US Death Certificates 2003–2023

**DOI:** 10.64898/2026.01.31.26345264

**Authors:** Michael Levitt, Ben Marten, Gal Oren, John P.A. Ioannidis

## Abstract

Death certificates record causes of death as reported by certifiers (Entity Axis) and as standardized by mortality coding rules (Record Axis). Conventional mortality statistics then reduce these to a single underlying cause, ignoring other contributing conditions; weighting schemes can instead distribute the mortality burden across all listed causes. We evaluated the impact of re-classification from Entity to Record axis and weighting across all 56,986,831 US death certificates from 2003–2023, mapping ICD-10 codes to 14 broad disease categories and testing three weighting schemes: W1 (50% weight to the underlying cause, 50% shared equally among contributing causes), W2 (equal weighting across all causes), and W2A (equal weighting at the ICD-10 code level). Entity and Record Axes agreed on underlying cause in 48,313,403 deaths (84.8%) by broad disease category and in 39,260,709 deaths (68.9%) by ICD-10 code (Supplementary Tables S5A, S5B); concordance reached 70.4% when using 3-character rather than 4-character ICD-10 categories. Re-classification markedly increased COVID-19 deaths (+92%) and Transport deaths (+43%) while markedly decreasing Other External Causes (-54%). Weighting substantially altered death burden attribution: COVID-19 (−44–63%) and Falls (−46–66%) decreased, Other External causes more than tripled (+204–254%), and deviations from unweighted counts were more pronounced with W2 and W2A than with W1. Weighting also brought disease-category counts closer to Entity Axis values and restored Respiratory seasonality patterns suppressed during the pandemic years. Systematic differences between reported and re-classified causes of death, and the choice of weighting scheme, profoundly alter disease burden estimates for several causes, with major implications for resource allocation and public health priorities.

**SIGNIFICANCE STATEMENT:** Standardized re-classification processes using mortality coding rules recast the selected causes of death in many death certificates. Moreover, vital statistics typically isolate a single underlying cause, while for many deaths multiple causes jointly lead to demise. Analysis of ∼57 million deaths in the USA (2003-2023) shows that a large proportion of deaths are re-classified by rules to different causes than those filled by original certifiers. Analyses that give weight not only to recorded underlying cause but also to other listed causes lead to markedly different estimates of deaths from several diseases. For example, the footprint of COVID-19 fatalities during the pandemic years decreases by 44-63%. Re-classification and weighting schemes may have profound impact on disease burden estimates and policy decisions.

## INTRODUCTION

Death certificates are widely known to have high rates of inaccuracies and inconsistencies (1-5), but they serve as the foundational data source for mortality statistics and for public health policies that depend on these statistics. Problems often arise at the level of filling out the certificate forms by physicians and other eligible health care practitioners who certify the causes of death. The certifiers may lack experience, make inadvertent errors, or have less than optimal knowledge of the complete death situation, circumstances, and past medical history of the deceased. Autopsies are uncommonly performed. To enhance standardization, for several decades now systems have been in place that process the crude reported death certificates and re-classify the causes of death with specific mortality coding rules (6,7). The certifier-entered information is termed Entity Axis, and the re-classified standardized information is termed Record Axis. The data of the Record Axis are the ones endorsed officially to represent the cleaned information on underlying causes of death and on other contributing causes. The mortality coding rules for the cleaning and re-classification process from the Entity to the Record Axis are laid out in detail (6,7). However, they are sometimes revised, when a new version of the International Classification of Diseases (ICD) is released and when new ICD codes are added or existing ones are modified. The introduction of COVID-19 coding in the ICD system during the pandemic was such an example of a prominent modification (8).

Even with the Record Axis standardization, major inaccuracies may persist if the original input at the Entity Axis is flawed in ways that cannot be amended by the re-classification rules. Studies of in-depth medical record audits demonstrate large discrepancies between death certificates and more appropriate determination of causes of death based on the in-depth assessments; this has been particularly prominent for deaths where multi-morbidity is involved such as COVID-19 (9-11). A pervasive issue relates to the fundamental traditional rule allowing only one underlying cause. In reality, many deaths have substantial contributions from multiple causes (12-22). The preference for listing a specific cause as underlying rather than contributing may change over time, depending on physician awareness and on how lethal that cause is perceived to be relative to competing causes (16). Evolving perceptions may create artefacts on the increase or decrease of specific diseases as underlying causes of death. Misclassifications of underlying and contributing causes become a greater challenge with increasing multimorbidity (17). For people with co-existing morbidities that have eroded their health, it may be best to give weight not just to one cause but many. Absent in-depth medical record audits, the listing of additional causes in the death certificates may provide some indication of the broader background for each death. Several authors have proposed weighting schemes where multiple contributing causes get a share of the death attribution (19-22).

Here we aimed to analyze all death certificates in the USA between 2003 and 2023 to empirically assess how common are changes between Entity and Record Axes and whether specific major patterns exist in these re-classifications. Moreover, we aimed to assess whether different weighting schemes for multiple causes of death may substantially affect the estimated burden of death from specific causes. We examined evolving patterns over time during this extended period, and separately in elderly and non-elderly populations paying attention to the pandemic period 2020-2023 and to COVID-19 deaths. COVID-19 represented a major change in cause of death classifications with extensive debate about the over- or under-counting of COVID-19 deaths (23).

## METHODS

### Death certificate data

Raw US death certificate data covering the 21-year period from 2003 to 2023 were obtained from the publicly accessible Centers for Disease Control and Prevention (CDC) National Center for Health Statistics (NCHS) FTP server (24). The dataset consists of annual mortality files (`mort2003us.zip` through `mort2023us.zip`). These archives were decompressed to yield fixed-width text files (`VS23MORT.DUSMCPUB`) containing death certificate data in specified columns. For each death, we extracted the complete information on ICD codes from both the Entity Axis and the Record Axis. We then classified the ICD codes into 14 broad disease categories, 7 natural ones (Circulatory, Cancer, Respiratory, Endocrine/Nutritional/Metabolic, Digestive, COVID-19, and Other Natural) and 7 external ones (Drug Poisoning, Transport, Suicide, Alcohol-induced, Falls, Homicide, Other External), following the classification scheme of Wrigley-Field et al (25) that simplifies a previously proposed grouping of the National Academies of Science, Engineering and Medicine (26). Our grouping is the same as in (25) except that we also have Falls as a separate broad category rather than merged with Other External, since reported Falls deaths have rapidly increased in the 21^st^ century and thus would be worth closer examination. For COVID-19 we used three ICD-10 codes (U07.1, U07.2, and U09.9) as done before (25). The few (n=518) occurrences of uncertain values (all were string “1”) under ICD-10 codes in the Record Axis were ignored.

When several ICD codes were mapped to the same broad disease category, they were usually counted only once in all analyses that used the broad disease categories (W2A is the exception). We used the broad-level classification to focus on major shifts across major groups of causes.

### Entity Axis versus Record Axis comparisons

We noted for each broad disease category how often it is listed as what might be assumed to be the underlying cause of death in the Entity Axis (i.e. appearing in the line in Part I of the death certificate filled out by certifying physician or other eligible health care practitioner) versus as underlying cause in the Record Axis. We also examined in each Axis for each broad disease category how often it was listed but not as underlying cause; how often it was listed at all (either as underlying cause or other); and how often it was listed as the only broad disease category in the death certificate. We then evaluated the underlying causes of death according to the two Axes and noted how often they agreed at the broad disease category level and at the specific ICD level. Focusing on the broad disease categories, we also recorded how often each cause was promoted to underlying cause of death from the re-classification of Entity Axis into Record Axis and how often it was demoted from underlying cause of death during that re-classification, the ratio of promotions to demotions, and the specific pairs of broad disease categories on which major shifts occurred. Specifically for COVID-19 we also examined the specific ICD categories involved in such promotion and demotion switches.

When there was more than one ICD code listed in the last line of Part I in the Entity Axis, we used the one listed last for the main analyses. As a sensitivity analysis, we examined the one listed first in the last line of Part I.

Of note, Entity Axis is not normally used for underlying cause reporting, therefore the comparison between Entity and Record Axis is done primarily to examine the effect of the re-classification process that generates the official cause data (Record Axis). Moreover, examination of the Entity Axis data allows to see whether weighted analyses considering multiple causes of death bring the death counts closer to the original Entity Axis entries.

### Weighting of multiple causes of death

We generated data for death counts comparing different weighting schemes using information from the Record Axis: the baseline W0 (counting only the listed underlying cause of death in the Record Axis); a hierarchical weighting scheme W1, where the underlying cause of death is given 50% of the weight, and the remaining contributing causes of death share equally among them the other 50% (e.g. if there are 2 contributing causes, then each of them gets 25% of the weight and the underlying cause gets 50% of the weight); an equal weighting scheme W2 where all causes share equally the weight (e.g. with two contributing causes, each of them gets 33.33% of the weight and the underlying cause also gets 33.33%) and a variant of W2 called W2A where all the causes (including repeated occurrences in the same broad category that are normally excluded) share the weight equally. Similar weighting schemes have been previously proposed in analyses of multiple cause of death data (19-22) and have used different levels of grouping of causes. Hence, we explored both the high-level broad categories grouping (W1, and W2) and the more granular ICD-level analyses (W2A). We estimated the absolute and relative magnitude of change between W1, W2, and W2A versus the baseline W0 (which represents the traditionally presented mortality statistics) for the number of deaths attributed to each of the broad death categories.

We mapped the baseline versus the W1, W2 and W2A estimates for each of the broad death categories per month in the 2003-2023 period. We also performed similar subgroup analyses for age groups of elderly (greater than 65 years old, GE65) and non-elderly (less than 65 years old, LT65). The plotted estimates were population-adjusted using annual mid-year US population estimates from CDC Wonder available through the Human Mortality Database (https://www.mortality.org/File/GetDocument/hmd.v6/USA/STATS/Population5.txt), with 2023 as the reference year. Scaling was performed separately for the overall population and for each age subgroup using overall and subgroup-specific population denominators, such that observed deaths were multiplied by the ratio of 2023 population to that year’s population within each stratum.

For the weighted analyses, we also performed two sensitivity analyses using the underlying cause information from the Record Axis and as additional causes the Part II causes listed from the Entity Axis; and using the underlying cause information from the Entity Axis (last line on Part I) and as additional causes the Part II causes listed in the Entity Axis. These sensitivity analyses aim to avoid the potential immediate causes of deaths (for example, sepsis) that might have otherwise contaminated the listing of other causes in the Record Axis.

### Software

All analyses were performed using Python scripts developed iteratively with LLM assistance (Claude, Anthropic). The LLM served as a coding assistant – the investigators specified the analytical requirements, reviewed all generated code for correctness, and validated results through multiple cross-checks including verifying that weighted totals sum to total deaths, comparing results across independent implementations, spot-checking calculations manually on subsets of data, and confirming concordance with published CDC statistics where available. The final Python scripts are deterministic and reproducible; they are provided along with the key data in https://github.com/scientific-computing-user/Reclassification-and-Weighting-of-Multiple-Causes-of-Death-US-Death-Certificates-2003-2023/.

## RESULTS

### Entity versus Record Axis

56,986,831 U.S. death certificates were issued in 2003–2023. Comparison of Entity and Record Axis data (Table 1) shows substantial changes for some broad categories of death such as COVID-19 and Other External causes. In relative terms, re-classification from Entity Axis to Record Axis almost doubles the number of COVID-19 deaths (+92%) and creates also major increases in the number of Falls deaths (+69%), Transport deaths (+43%), and modest increases in the number of Suicide (+25%), Homicide (+30%), Endocrine (+16%) and Cancer deaths (+12%). Re-classification leaves Circulatory, Digestive, Alcohol-related and Drug Poisoning deaths largely unchanged. Conversely, it causes major decreases in Other External causes (-54%), and modest decreases in Other Natural causes (-14%), and Respiratory deaths (-11%).

**Table 1.**
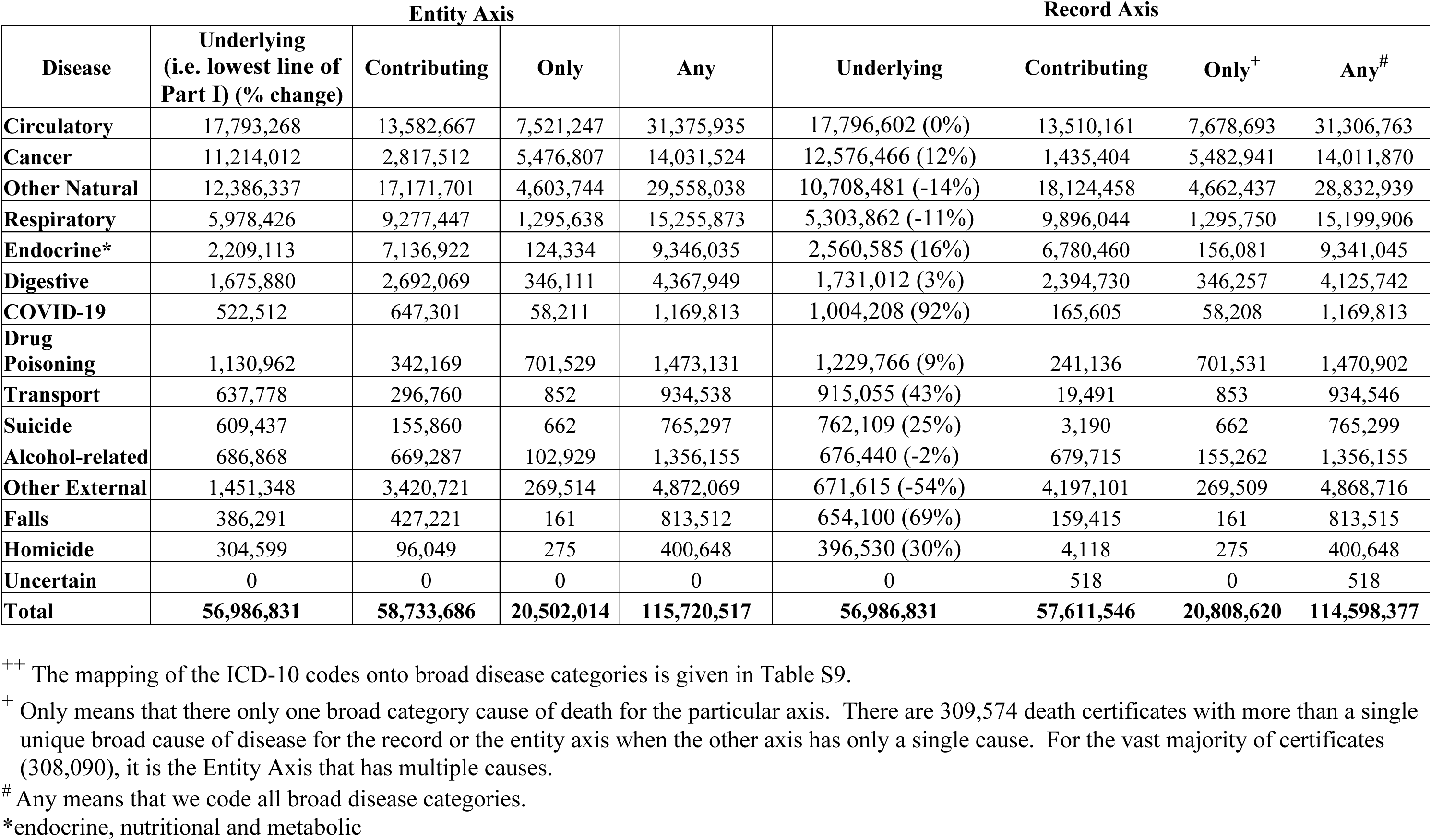
Causes of death classified in broad disease categories^++^ in Entity Axis and Record Axis, USA 2003-2023 (when there was more than one ICD code listed in the last line of Part I in the Entity Axis, we used the one listed last).

The availability of additional contributing causes of death more than doubles the total number of available causes (n=57,611,546 more) even with the clustering in broad categories. For both Axes, slightly over a third of the deaths (n=20,502,014 or 20,808,620) have only one of the broad disease categories represented in the death certificate. The relative ranking changes substantially for some categories such as COVID-19 and Cancer when any cause is considered instead of only the underlying cause. There is a large diversity across the broad categories on the ratio of their appearance as contributing causes rather than as underlying causes (ranging from near zero to 6.25, see Supplementary Table S1).

In 7,749,865 (13.6%) of the 56,986,831 deaths, the last line of Part I in the death certificate had multiple ICD codes listed. When assuming as underlying cause of death the first ICD listed in the last line of Part I instead of the last ICD listed in that line, the respective results and changes versus the main analysis are shown in Supplementary Tables S2 and S3.

Table 2 shows the specific transitions across the broad disease categories between the Entity Axis and the Record Axis for the underlying cause of death. Entity and Record Axes agreed on underlying cause (the Entity “underlying cause” is that reported in the lowest, last line of Part I) in 48,313,403 (84.8%) of the total deaths based on broad disease categories and in 39,260,709 (68.9%) based on ICD-10 codes (Supplementary Table S5A) and in 30,806,674 (70.4%) when we used 3-character ICD-10 categories instead of 4-characters (e.g. J44 instead of J449). Concordance was 85.7% and 70.1%, respectively, in pre-pandemic years (2003-2019) and 81.6% and 65.0%, respectively, during pandemic years (2020-2023). Discordance rates slowly increased between 2003 and 2019 (by about 15%) and then had a marked increase in 2020 and 2021 and partial recovery in 2022-2023.

**Table 2.** Transition matrix from Entity Axis (columns) to Record Axis (rows). When there was more than one ICD code listed in the last line of Part I in the Entity Axis, we used the one listed last.

| **ENTITY** | **Circu**<br>**latory** | **Cancer** | **Other**<br>**Natural** | **Respi**<br>**ratory** | **Endoc**<br>**rine\*** | **Digestive** | **COVID**<br>**-19** | **Drug**<br>**Poiso**<br>**ning** | **Trans**<br>**port** | **Suicide** | **Alcohol** | **Other**<br>**Exter**<br>**nal** | **Falls** | **Homi**<br>**cide** | **Total**<br>**Record**<br>**Axis** |
| --- | --- | --- | --- | --- | --- | --- | --- | --- | --- | --- | --- | --- | --- | --- | --- |
| **RECORD** |  |  |  |  |  |  |  |  |  |  |  |  |  |  |  |
| **Circulatory** | 15,936,312 | 61,223 | 1,092,514 | 408,494 | 143,301 | 49,031 | 7,330 | 16,246 | 1,313 | 104 | 12,981 | 63,365 | 4,047 | 341 | **17,796,602** |
| **Cancer** | 344,036 | 11,020,772 | 598,117 | 360,314 | 109,320 | 104,079 | 3,591 | 1,323 | 135 | 26 | 14,196 | 19,895 | 640 | 22 | **12,576,466** |
| **Other Natural** | 628,963 | 53,307 | 9,350,816 | 406,374 | 130,961 | 60,841 | 4,473 | 7,028 | 714 | 68 | 16,553 | 45,311 | 2,840 | 232 | **10,708,481** |
| **Respiratory** | 297,054 | 29,430 | 506,545 | 4,346,838 | 57,638 | 17,118 | 2,539 | 2,832 | 239 | 14 | 4,870 | 37,738 | 966 | 41 | **5,303,862** |
| **Endocrine\*** | 379,340 | 13,610 | 364,998 | 60,867 | 1,711,842 | 11,766 | 1,924 | 2,013 | 111 | 8 | 4,529 | 9,227 | 329 | 21 | **2,560,585** |
| **Digestive** | 68,911 | 14,182 | 187,179 | 54,731 | 15,488 | 1,372,198 | 338 | 1,017 | 159 | 8 | 2,568 | 13,926 | 257 | 50 | **1,731,012** |
| **COVID-19** | 54,475 | 10,168 | 119,181 | 288,936 | 21,328 | 5,017 | 501,747 | 204 | 48 | 5 | 567 | 2,251 | 271 | 10 | **1,004,208** |
| **Drug Poisoning** | 16,732 | 1,578 | 13,339 | 7,380 | 2,855 | 1,906 | 125 | 1,088,054 | 96 | 482 | 87,860 | 9,181 | 83 | 95 | **1,229,766** |
| **Transport** | 4,014 | 169 | 3,570 | 1,686 | 455 | 242 | 26 | 1,086 | 634,750 | 1 | 2,378 | 266,638 | 31 | 9 | **915,055** |
| **Suicide** | 531 | 443 | 9,809 | 359 | 121 | 78 | 14 | 1,943 | 5 | 608,669 | 966 | 139,167 | 3 | 1 | **762,109** |
| **Alcohol-related** | 17,325 | 2,231 | 48,391 | 11,891 | 8,062 | 48,038 | 87 | 3,809 | 50 | 31 | 530,942 | 5,339 | 233 | 11 | **676,440** |
| **Other External** | 29,504 | 4,740 | 63,407 | 22,983 | 5,584 | 4,561 | 89 | 4,721 | 107 | 14 | 4,980 | 530,516 | 391 | 18 | **671,615** |
| **Falls** | 15,552 | 2,144 | 27,326 | 7,253 | 1,974 | 929 | 229 | 426 | 39 | 2 | 3,402 | 218,625 | 376,199 | 0 | **654,100** |
| **Homicide** | 519 | 15 | 1,145 | 320 | 184 | 76 | 0 | 260 | 12 | 5 | 76 | 90,169 | 1 | 303,748 | **396,530** |
| **Total Entity Axis** | **17,793,268** | **11,214,012** | **12,386,337** | **5,978,426** | **2,209,113** | **1,675,880** | **522,512** | **1,130,962** | **637,778** | **609,437** | **686,868** | **1,451,348** | **386,291** | **304,599** | **56,986,831** |

\*endocrine, nutritional and metabolic

In sensitivity analysis using the first ICD listed in the last line of Part I instead of the last listed one, concordance rates were slightly lower (82.3% for broad category and 67.7% for ICD level) (see Supplementary Table S5A) and 69.1% with ICD-10 at 3-character categories.

Focusing on the broad death categories in Table 2 that have the most striking relative increases (COVID-19, Transport) or relative decreases (Other External, Alcohol) shows that a few specific category transitions are mostly responsible for the changes. For COVID-19, 288,936 deaths (29% of the 1,004,208 total) in the Record Axis are imported from the Respiratory category in the Entity Axis and another 119,181 deaths (12%) are imported from the Other Natural deaths in the Entity Axis. Transport deaths in the Record Axis receive 266,638 deaths (29% of the 915,055 total in the Record Axis) from the Other External causes in the Entity Axis. Other External causes get often re-classified also as Falls (218,625, 33% of Falls total deaths of 654,100 in Record Axis) and Suicide receives 139,167 deaths (18.2% of its total of 762,109 in the Record Axis) from Other External causes in the Entity Axis. Alcohol deaths in the Entity Axis often get re-classified as Drug Poisoning (87,860 deaths) in the Record Axis.

For COVID-19 deaths, the number of promotions to underlying cause of death (n=502,461) outnumber demotions (n=20,765) by 24-fold in the Entity to Record Axis re-classifications. Promotions largely re-classify the underlying cause of death away from respiratory and infection syndromes (Table S4). Most prominently, the ICD-10 J189 code of pneumonia represents 43.8% of promotions. The demotions are much rarer and shift the underlying cause of death toward very diverse chronic disease causes.

### Weighted causes of death

Table 3, panel 1 shows the unweighted (W0) and W1-, W2-, and W2A-weighted counts for deaths in broad disease categories for 2003-2023 and separately for the pandemic years 2020-2023 and Table 4 does the same separately for age strata. The weighting results in major changes for the relative contribution of some disease categories to the overall death burden and usually changes are more prominent with W2 and W2A.

**Table 3.** Underlying death counts for broad disease categories for 2003-2023 and for the pandemic years 2020-2023 with different weighting schemes (W0: no weighting, only the underlying cause is counted; W1: 50% weight for the underlying cause and the other 50% shared equally among other causes; W2: weight shared equally among all causes; W2A: weight shared equally among all causes but considering ICD-level causes rather than just broad categories). The upper panel weights the counts of ICD on the Record Axis. The middle panel weights the counts for the underlying ICD on the Record Axis with the contributing ICD on the Entity Axis. The lower panel weights the counts for the underlying ICD on the Entity Axis (last line of Part I) with the contributing ICD on the Entity Axis.

| Counts and percentage relative to W0 for the counts of ICD causes from the Record Axis |  |  |  |  |  |  |  |  |
| --- | --- | --- | --- | --- | --- | --- | --- | --- |
|  | All Years (2003-2023) |  |  |  | Pandemic Years (2020-2023) |  |  |  |
| Disease | W0 | W1 | W2 | W2A | W0 | W1 | W2 | W2A |
| Circulatory | 17,797 | 17,960 (+1) | 18,060 (+1) | 17,045 (-4) | 3,715 | 3,867 (+4) | 3,942 (+6) | 3,688 (-1) |
| Cancer | 12,576 | 9,392 (-25) | 8,546 (-32) | 8,832 (-30) | 2,494 | 1,827 (-27) | 1,615 (-35) | 1,678 (-33) |
| Other Natural | 10,708 | 13,538 (+26) | 14,424 (+35) | 14,186 (+32) | 2,468 | 3,131 (+27) | 3,378 (+37) | 3,286 (+33) |
| Respiratory | 5,304 | 6,122 (+15) | 6,287 (+19) | 6,481 (+22) | 1,046 | 1,388 (+33) | 1,497 (+43) | 1,511 (+44) |
| Endocrine* | 2,561 | 2,653 (+4) | 2,707 (+6) | 3,404 (+33) | 679 | 733 (+8) | 766 (+13) | 950 (+40) |
| Digestive | 1,731 | 1,636 (-5) | 1,565 (-10) | 1,668 (-4) | 394 | 365 (-7) | 347 (-12) | 375 (-5) |
| COVID-19 | 1,004 | 566 (-44) | 370 (-63) | 441 (-56) | 1,004 | 566 (-44) | 370 (-63) | 441 (-56) |
| Drug Poisoning | 1,230 | 1,140 (-7) | 1,087 (-12) | 988 (-20) | 426 | 396 (-7) | 377 (-11) | 342 (-20) |
| Transport | 915 | 461 (-50) | 380 (-58) | 437 (-52) | 192 | 97 (-50) | 81 (-58) | 91 (-52) |
| Suicide | 762 | 382 (-50) | 308 (-60) | 363 (-52) | 174 | 87 (-50) | 69 (-60) | 83 (-52) |
| Alcohol-related | 676 | 547 (-19) | 512 (-24) | 574 (-15) | 202 | 162 (-20) | 150 (-26) | 170 (-16) |
| Other External | 672 | 2,039 (+204) | 2,376 (+254) | 2,104 (+213) | 145 | 454 (+212) | 533 (+266) | 482 (+231) |
| Falls | 654 | 351 (-46) | 225 (-66) | 269 (-59) | 180 | 97 (-46) | 59 (-67) | 72 (-60) |
| Homicide | 397 | 200 (-50) | 140 (-65) | 195 (-51) | 97 | 49 (-50) | 34 (-66) | 48 (-51) |

|  | All Years (2003-2023) |  |  |  | Pandemic Years (2020-2023) |  |  |  |
| --- | --- | --- | --- | --- | --- | --- | --- | --- |
| Disease | W0 | W1 | W2 | W2A | W0 | W1 | W2 | W2A |
| Circulatory | 17,816<sup>+</sup> | 16,679 (-6) | 16,594 (-7) | 16,330 (-8) | 3,715 | 3,556 (-4) | 3,568 (-4) | 3,484 (-6) |
| Cancer | 12,576 | 10,789 (-14) | 10,396 (-17) | 10,514 (-16) | 2,494 | 2,085 (-16) | 1,971 (-21) | 2,004 (-20) |
| Other Natural | 10,692 | 13,742 (+29) | 14,228 (+33) | 13,961 (+31) | 2,468 | 3,191 (+29) | 3,336 (+35) | 3,250 (+32) |
| Respiratory | 5,308 | 4,794 (-10) | 4,694 (-12) | 4,880 (-8) | 1,046 | 965 (-8) | 952 (-9) | 1,000 (-4) |
| Endocrine* | 2,550 | 3,448 (+35) | 3,706 (+45) | 3,865 (+52) | 679 | 920 (+35) | 1,002 (+47) | 1,046 (+54) |
| Digestive | 1,731 | 1,662 (-4) | 1,641 (-5) | 1,695 (-2) | 394 | 378 (-4) | 372 (-6) | 388 (-1) |
| COVID-19 | 1,004 | 784 (-22) | 705 (-30) | 733 (-27) | 1,004 | 784 (-22) | 705 (-30) | 733 (-27) |
| Drug Poisoning | 1,230 | 1,165 (-5) | 1,142 (-7) | 1,099 (-11) | 426 | 403 (-5) | 394 (-7) | 380 (-11) |
| Transport | 915 | 852 (-7) | 839 (-8) | 845 (-8) | 192 | 178 (-8) | 174 (-9) | 176 (-9) |
| Suicide | 762 | 444 (-42) | 428 (-44) | 431 (-43) | 174 | 95 (-46) | 91 (-48) | 92 (-47) |
| Alcohol-related | 680 | 665 (-2) | 655 (-4) | 677 (-0) | 202 | 192 (-5) | 187 (-8) | 195 (-4) |
| Other External | 672 | 1,246 (+85) | 1,306 (+94) | 1,273 (+90) | 145 | 295 (+103) | 310 (+113) | 305 (+110) |
| Falls | 654 | 478 (-27) | 420 (-36) | 445 (-32) | 180 | 125 (-31) | 105 (-42) | 114 (-37) |
| Homicide | 397 | 240 (-40) | 235 (-41) | 237 (-40) | 97 | 53 (-46) | 52 (-47) | 52 (-46) |

|  | All Years (2003-2023) | Pandemic Years (2020-2023) |
| --- | --- | --- |

| Disease | W0 | W1 | W2 | W2A | W0 | W1 | W2 | W2A |
| --- | --- | --- | --- | --- | --- | --- | --- | --- |
| **Circulatory** | 17,793 | 16,726 (-6) | 16,670 (-6) | 16,386 (-8) | 3,773 | 3,618 (-4) | 3,631 (-4) | 3,541 (-6) |
| **Cancer** | 11,214 | 9,798 (-13) | 9,484 (-15) | 9,593 (-14) | 2,208 | 1,882 (-15) | 1,790 (-19) | 1,821 (-18) |
| **Other Natural** | 12,386 | 14,917 (+20) | 15,271 (+23) | 14,997 (+21) | 2,960 | 3,525 (+19) | 3,623 (+22) | 3,536 (+19) |
| **Respiratory** | 5,978 | 5,224 (-13) | 5,049 (-16) | 5,259 (-12) | 1,392 | 1,206 (-13) | 1,157 (-17) | 1,215 (-13) |
| **Endocrine\*** | 2,209 | 3,225 (+46) | 3,528 (+60) | 3,713 (+68) | 598 | 873 (+46) | 969 (+62) | 1,021 (+71) |
| **Digestive** | 1,676 | 1,627 (-3) | 1,615 (-4) | 1,669 (-0) | 388 | 374 (-4) | 370 (-5) | 386 (-0) |
| **COVID-19** | 523 | 422 (-19) | 382 (-27) | 401 (-23) | 523 | 422 (-19) | 382 (-27) | 401 (-23) |
| **Drug Poisoning** | 1,131 | 1,120 (-1) | 1,109 (-2) | 1,072 (-5) | 389 | 387 (-1) | 382 (-2) | 370 (-5) |
| **Transport** | 638 | 653 (+2) | 645 (+1) | 651 (+2) | 142 | 140 (-2) | 138 (-3) | 139 (-2) |
| **Suicide** | 609 | 371 (-39) | 359 (-41) | 362 (-41) | 140 | 82 (-41) | 79 (-44) | 80 (-43) |
| **Alcohol-related** | 687 | 660 (-4) | 650 (-5) | 670 (-2) | 208 | 190 (-9) | 185 (-11) | 192 (-8) |
| **Other External** | 1,451 | 1,703 (+17) | 1,709 (+18) | 1,673 (+15) | 307 | 385 (+25) | 386 (+26) | 382 (+24) |
| **Falls** | 386 | 350 (-9) | 330 (-15) | 351 (-9) | 111 | 92 (-17) | 84 (-25) | 91 (-18) |
| **Homicide** | 305 | 190 (-38) | 187 (-38) | 189 (-38) | 78 | 44 (-44) | 43 (-45) | 43 (-44) |

\*endocrine, nutritional metabolic
<sup>†</sup>There are small differences in W0 counts with the different classification scheme used in this panel.

**Table 4.** Death counts for broad disease categories for 2003-2023 and for the pandemic years 2020-2023 with different weighting schemes (W0: no weighting, only the underlying cause is counted; W1: 50% weight for the underlying cause and the other 50% shared equally among other causes; W2: weight shared equally among all causes; W2A: weight shared equally among all causes but considering ICD-level causes rather than just broad categories) separately for deaths under 65 years old and for deaths 65 years old or older. The causes weighted are all on the Record Axis.

**Death counts for non-elderly, under 65 years old, in thousands (% of W0)**
| Disease | All Years (2003-2023) |  |  |  | Pandemic Years (2020-2023) |  |  |  |
| --- | --- | --- | --- | --- | --- | --- | --- | --- |
|  | W0 | W1 | W2 | W2A | W0 | W1 | W2 | W2A |
| **Circulatory** | 3,312 | 3,396 (+3) | 3,425 (+3) | 3,288 (-1) | 695 | 721 (+4) | 734 (+6) | 703 (+1) |
| **Cancer** | 3,635 | 2,744 (-25) | 2,524 (-31) | 2,576 (-29) | 638 | 469 (-27) | 417 (-35) | 428 (-33) |
| **Other Natural** | 2,086 | 2,890 (+39) | 3,136 (+50) | 3,068 (+47) | 402 | 603 (+50) | 678 (+69) | 654 (+63) |
| **Respiratory** | 808 | 1,074 (+33) | 1,147 (+42) | 1,181 (+46) | 168 | 267 (+59) | 302 (+79) | 302 (+79) |
| **Endocrine\*** | 721 | 715 (-1) | 713 (-1) | 858 (+19) | 180 | 189 (+5) | 195 (+8) | 232 (+29) |
| **Digestive** | 532 | 561 (+5) | 557 (+5) | 579 (+9) | 116 | 117 (+1) | 115 (-1) | 122 (+5) |
| **COVID-19** | 248 | 138 (-44) | 89 (-64) | 106 (-57) | 248 | 138 (-44) | 89 (-64) | 106 (-57) |
| **Drug Poisoning** | 1,159 | 1,076 (-7) | 1,027 (-11) | 934 (-19) | 396 | 368 (-7) | 351 (-11) | 318 (-20) |
| **Transport** | 745 | 374 (-50) | 311 (-58) | 358 (-52) | 153 | 77 (-50) | 65 (-57) | 73 (-52) |
| **Suicide** | 622 | 312 (-50) | 254 (-59) | 297 (-52) | 139 | 69 (-50) | 56 (-60) | 66 (-52) |
| **Alcohol-related** | 526 | 426 (-19) | 402 (-24) | 450 (-15) | 152 | 122 (-20) | 113 (-25) | 128 (-16) |
| **Other External** | 356 | 1,278 (+259) | 1,472 (+314) | 1,303 (+267) | 74 | 278 (+274) | 322 (+333) | 290 (+290) |
| **Falls** | 100 | 52 (-48) | 35 (-65) | 42 (-58) | 23 | 12 (-48) | 8 (-65) | 9 (-59) |
| **Homicide** | 376 | 189 (-50) | 132 (-65) | 185 (-51) | 92 | 46 (-50) | 32 (-66) | 46 (-51) |

**Death counts for elderly, 65 years old or older, in thousands (% of W0)**
| Disease | All Years (2003-2023) |  |  |  | Pandemic Years (2020-2023) |  |  |  |
| --- | --- | --- | --- | --- | --- | --- | --- | --- |
|  | W0 | W1 | W2 | W2A | W0 | W1 | W2 | W2A |
| **Circulatory** | 14,485 | 14,564 (+1) | 14,635 (+1) | 13,757 (-5) | 3,021 | 3,147 (+4) | 3,208 (+6) | 2,986 (-1) |
| **Cancer** | 8,942 | 6,648 (-26) | 6,022 (-33) | 6,257 (-30) | 1,855 | 1,358 (-27) | 1,197 (-35) | 1,251 (-33) |
| **Other Natural** | 8,623 | 10,648 (+23) | 11,287 (+31) | 11,119 (+29) | 2,066 | 2,528 (+22) | 2,701 (+31) | 2,633 (+27) |
| **Respiratory** | 4,496 | 5,048 (+12) | 5,140 (+14) | 5,299 (+18) | 878 | 1,121 (+28) | 1,196 (+36) | 1,210 (+38) |
| **Endocrine\*** | 1,839 | 1,938 (+5) | 1,994 (+8) | 2,546 (+38) | 499 | 544 (+9) | 571 (+14) | 718 (+44) |
| **Digestive** | 1,199 | 1,076 (-10) | 1,008 (-16) | 1,089 (-9) | 278 | 248 (-11) | 232 (-17) | 253 (-9) |
| **COVID-19** | 756 | 427 (-43) | 281 (-63) | 334 (-56) | 756 | 427 (-43) | 281 (-63) | 334 (-56) |
| **Drug Poisoning** | 70 | 64 (-9) | 60 (-15) | 54 (-23) | 30 | 28 (-8) | 26 (-13) | 24 (-21) |
| **Transport** | 171 | 87 (-49) | 69 (-60) | 79 (-54) | 39 | 20 (-49) | 16 (-60) | 18 (-54) |
| **Suicide** | 140 | 70 (-50) | 54 (-62) | 66 (-53) | 36 | 18 (-50) | 13 (-62) | 17 (-53) |
| **Alcohol-related** | 150 | 121 (-20) | 110 (-27) | 124 (-17) | 51 | 40 (-20) | 37 (-28) | 42 (-17) |
| **Other External** | 316 | 760 (+141) | 904 (+186) | 801 (+153) | 71 | 176 (+148) | 210 (+196) | 192 (+171) |
| **Falls** | 554 | 299 (-46) | 190 (-66) | 227 (-59) | 158 | 85 (-46) | 51 (-67) | 62 (-61) |
| **Homicide<sup>+</sup>** | 20 | 11 (-48) | 8 (-63) | 10 (-51) | 5 | 3 (-49) | 2 (-64) | 3 (-52) |

\*endocrine, nutritional metabolic
<sup>+</sup>The percentage changes using the complete counts not those rounded in thousands.

With weighting, COVID-19 (-44-63%), Transport (-50-58%), Suicide (-50-60%), Falls (-46-66%) and Homicide (-50-65%) death counts decrease drastically and there are also major 19-32% decreases for Cancer and Alcohol-related deaths. Conversely, Other External causes more than triple (+204-254%); Other Natural, Respiratory and Endocrine, Nutrition and Metabolic deaths increase by 4-35% in the 2003-2023 data and slightly more prominently (8-44%) in the pandemic years alone. There are small decreases in Drug Poisoning and Digestive deaths, while Circulatory deaths remain largely unchanged. The rank order of causes changes substantially in the weighted analyses. For all years, all three weighting schemes promote Other External causes from rank 12 to 6. At the same time Digestive, COVID-19, Drug Poisoning, Suicide and Transport causes are demoted by between 1 and 3 ranks.

Table 3, panel 2 shows the weighted analyses using the Record Axis underlying cause with contribution causes from Part II of the Entity Axis instead of the causes from Record Axis as presented above: results were largely similar. Moreover, Figure S1 shows how the percent changes to each broad disease category for the different weighting schemes (W1, W2 and W2A) are highly correlated with correlation coefficients for the entire period of 0.809, and of 0.824 for the pandemic years) for calculations done using the causes on Part II of the Entity Axis instead of causes on the Record Axis. Table 3, panel 3 shows that the results were largely similar when only information from Entity Axis (Entity underlying cause from last line of Part I with contributing causes from Part II) was used in the weighting.

These patterns follow directly from the disparity in count between underlying and contributing causes of death already seen in Table 1 and Supplementary Table S1. If there are equal counts of the two categories, sensible weighting is expected to have little effect (e.g. Circulatory). If there are fewer counts in contributing causes of death, such weighting decreases the death counts (e.g. COVID-19, Transport, Suicide, Falls) and if there are more counts in contributing causes of death, such weighting increases the death counts (e.g. Respiratory, Endocrine, Other External). In the extreme increases with weighting that are seen for Other External causes, the key reasons are respiratory tract obstruction, fractures, and injuries (Supplementary Table S6), and many of these ICD-10 codes probably reflect the nature rather than the cause of the injury.

Among deaths in ages under 65 years (Table 4), Cancer deaths are the most common cause in the unweighted counts, but with weighting the Circulatory category becomes the most common one. With the weighting schemes W2 and W2A Cancer gets to third rank after Circulatory and Other Natural causes. Conversely, with weighting the relative increases in Respiratory deaths and deaths due to Other Natural causes are more prominent in this age stratum, especially in the pandemic years. With weights W2 and W2A COVID-19 deaths are cut the most (by 64% and 57%, respectively) in this age stratum. In the age stratum of age 65 years and older (also in Table 4), for most of the broad death categories the relative change in death counts with weighting are similar as in the non-elderly stratum. However, there are some exceptions. Boosting of Other External causes is less pronounced while boosting of Respiratory, Endocrine and Other Natural causes is more pronounced than in the non-elderly.

The corrections to the Record Axis underlying death counts made by all three weighting schemes are such as to make the counts become more similar to what they were in the Entity Axis of the death certificate. Specifically, the Pearson correlation coefficients of the fractional changes caused by weighting and the fractional change between Entity and Record axes is between 0.96 and 0.97 (Supplementary Table S7).

Figure 2 shows the evolution of death numbers between 2003 and 2023 for each of the broad disease categories using the unweighted W0 death counts and the W1-weighted death counts. There is observed seasonality for several broad categories. For Respiratory, but also for Circulatory, Endocrine Nutrition and Metabolic, Falls, and Other Natural disease causes there are annual peaks in the colder seasons and troughs in the summer, while for Transport and Suicide deaths the opposite seasonality pattern is seen. In the baseline W0 counts, the seasonality is largely lost for Respiratory deaths during the pandemic years, but it is recovered with the weighting. Supplementary Figures S3 and S4 show separate data for the elderly and non-elderly age groups. Similar patterns are seen with W2 and W2A weights (Supplementary Figures S5 and S6, and age subgroups in Supplementary Figures S7, S8, S9, and S10), although the magnitude of the deviations from W0 tend to be bigger with W2 and W2A than with W1.

**Figure 1.**
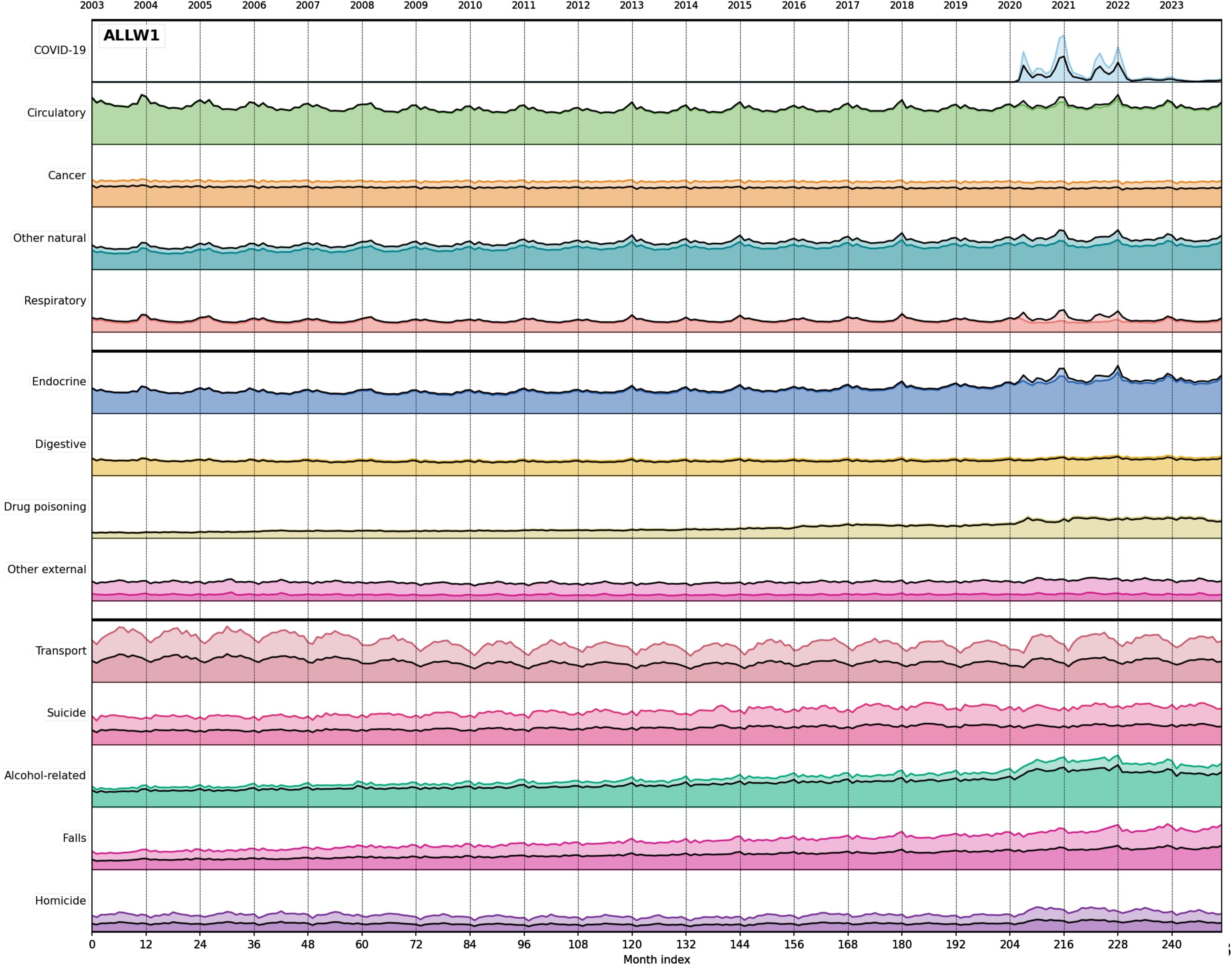
Population-normalized monthly deaths across 14 broad categories during 2003-2023 for all ages comparing unweighted W0 with W1 weighting. In this figure as well as in supplementary figures S3 to S10, unweighted is a colored line and weighted is a thick black line. Disease categories are grouped by fixed Y-axis scales to allow visual comparison across panels. The top band (COVID-19, Circulatory, Cancer, Other natural, Respiratory) uses 0-130,000 deaths/month; the middle band (Endocrine, Digestive, Drug poisoning, Other external) uses 0-25,000; the bottom band (Transport, Suicide, Alcohol-related, Falls, Homicide) uses 0-6,000. All values are population-scaled to 2023 levels.

There are also robust linear and logarithmic increases over time for some categories and age-groups (Supplementary Table S8; Supplementary Figure S11), such as Drug Poisoning (linear trend 6.7% per year), Alcohol (4.7% per year) and Falls (4.3% per year) deaths and less prominently for Other Natural (1.9% per year) and Suicide (2.0% per year). For the elderly, there are gradual decreases over time for Circulatory (-2.5% per year) and Cancer (-1.9% per year for ages 65 or over) deaths. Linear and logarithmic annual percentage changes are very similar and weighting has little clear effect on these rates of change.

## DISCUSSION

Our analysis of almost 57 million death certificates in the USA over 21 years offers insights on the impact of re-classifications during processing and of weighting for multimorbidity with multiple causes rather than a single death cause shaping each fatality. One third of deaths change ICD code for the selected underlying cause upon processing from the crude reported certificate to the version used in official national statistics and about half of them involve a shift even to a different broad disease category. Shifts have become more common over time and substantially escalated in the pandemic years, especially in 2020-2021, with the introduction of COVID-19 coding. Re-classification almost doubled the number of deaths where COVID-19 was selected as the underlying cause. Conversely, consideration of multimorbidity with weighting for multiple causes of death cut about half to two-thirds of the COVID-19 burden. It also generated substantive changes in the absolute death counts and in the relative ranking of several other causes of death. Weighting brought again the death counts for different disease categories closer to the Entity Axis. Seasonality peaks for Respiratory deaths were affected by the use or not of weighting.

The relatively high, and increasing, rates of disagreement between Record and Entity Axis are worrisome. They may reflect a high, and increasing, level of inaccuracy in filling death certificates. The escalating lack of concordance during 2020-2021 may reflect lack of acquaintance of certifiers in the new COVID-19 coding; or a particular difficulty in describing COVID-19 deaths and properly assigning the underlying cause of death in the standard framework where only one underlying cause is allowed. COVID-19 is a classic example where almost all deaths occurred in the presence of major co-morbidities, and where several co-morbidities had strong associations with fatal outcomes in SARS-CoV-2 infections (27-29). It is thus possible that the WHO rules which were embedded also in the USA re-classification mortality coding rules (much like in other countries) were markedly overestimating COVID-19 death burden (23). Record Axis COVID-19 deaths have included many deaths that had multiple other causes with substantive contribution to the fatal outcome. Even beyond COVID-19, deaths with high levels of multimorbidity are probably becoming more common over time posing a challenge for certifiers to select a single underlying cause. Additional analyses using weighting for multiple causes of deaths need to be conducted across diverse countries, with priority on those that have the most complete information from death certificates. Some of our findings may be more specific to the USA (e.g. drug use and homicide counts) while others, such as changes in recording and coding of COVID-19 were experienced in many countries around the world.

Specific non-COVID-19 re-classifications are heavily represented, especially those involving the shift from Other External causes to Transport, Suicide, and Falls deaths. Many such shifts may be appropriate to capture the more precise initiating event that led to death. Some re-classifications may be debatable, however. For example, Alcohol-related death counts were overall reduced by the re-classification, but this may be underestimating the contribution of alcohol to many other conditions that eventually kill people. Moreover, it is well documented that alcohol is already grossly under-reported in death certificates: for example, traffic census data show that in motor traffic crash deaths with detected high alcohol levels, only 16% of death certificates in the USA mention alcohol, with wide variation across states (2-81%) (30).

The weighting schemes attempt to correct for the multi-factorial nature of deaths, a scenario that has become extremely common, largely the norm rather than the exception, in the 21^st^ century. The paradigm of listing only one underlying cause of death was quite appropriate for eras where previously healthy, young people were dying from acute infectious diseases or violence. In the current circumstances, this paradigm has very questionable validity. Most people die from the concerted impact of multiple causes that undermine their health over a variable range of time. Some causes act on a shorter time horizon than others. The exact extent to which causes other than the underlying cause should be considered is debated. We tried three weighting options inspired by previous work (19-22), extending them to consider other causes both at the level of broad categories and at ICD level.

The weighted analyses yielded largely similar results in terms of direction, although W2 and W2A tend to produce larger deviations from the baseline unweighted W0 than W1. Weighting tends to bring the corrected death counts closer to what certifiers had selected in the Entity Axis. When multimorbidity is extreme (like in COVID-19), the more prominently altered estimates may be more appropriate. A much lower than officially recorded number of COVID-19 deaths is commensurate also with the small loss of life years estimated for many of those fatalities (31). It also agrees with the findings of several in-depth audit evaluations of medical records in different countries where large percentages of previously officially proclaimed COVID-19 deaths are found to have other primary, underlying death causes (9-11). It is thus likely that the true pandemic burden of COVID-19 corresponded to only less than half or even just a third of the officially assigned COVID-19 deaths in the USA. Probably COVID-19 during 2020-2023 accounted cumulatively for ∼100,000 deaths among people less than 65 years old, and ∼300,000 deaths among people 65 years old and older.

Changes in the frequency of specific death causes over time may reflect genuine changes due to changing incidence (e.g. infectious disease waves) or improvements in diagnostic and therapeutic management and outcomes. However, changes may also reflect an alteration of the perceived importance of a death cause among health care practitioners and in the community in general. This may affect whether a cause is listed at all and whether it is chosen to be listed as underlying or contributing. An example has been dissected in the case of Alzheimer’s disease where an increase in its listing as underlying cause was accompanied by a decrease in its listing as contributing cause (16). In our data, perception issues may underlie the observed patterns for COVID-19. Apparently, in the very early phase of the pandemic, some COVID-19 deaths were not recorded, either because their etiology was entirely missed or because certifiers may have not considered COVID-19 as the underlying cause. At this early phase, substantial under-reporting of COVID-19 deaths may have been reflected in increased death rates from other listed causes (32). Later the inverse pattern probably predominated, with severe over-counting of COVID-19 deaths in an environment where the pandemic was a dominant pervasive theme in health, media and in society at large, excessive testing was performed (23).

Perception, testing, and recognition issues are most stark with COVID-19 but may affect also in more subtle and chronic ways several major causes of deaths from chronic disease. For example, other investigators have noted that perception issues may propel ischemic heart disease, rather than diabetes or hypertension or heart failure to be listed as the underlying cause of death (22). Weighting of multiple death causes may correct some of this injustice of all-or-none attribution. However, it has also been argued that weighting may be inappropriate for some causes that might be truly the main and only determinant of fatality, as for example for many cancers (22). Moreover, some additional listed external causes, such as injuries and fractures, and respiratory tract obstruction may reflect mostly the nature of an injury and death process rather than causes that should get a share of the weight. Furthermore, one could further question whether the chronic trends (with or without weighting) reflect real changes or perception changes. This dilemma could affect even the most common causes of disease, i.e. circulatory and cancer deaths. Certifiers may be certifying causes according to the increase or decrease of the reputational presence of each disease in the environment where they practice. Perception biases could make death certificate data become self-fulfilling prophecies about the burden of specific diseases and the evolution of this burden over time.

Our work has several limitations. First, it should be acknowledged that the selected grouping of death causes can make a difference. Here we used primarily high-level, broad categories of death, to avoid the frequent dilemmas between different ICD-10 codes within the same system, as in ischemic heart disease and hypertension (19-22). This approach captures only the most prominent shifts in causal categorization. At the level of single ICD-10 codes, shifts both with re-classification and with weighting were documented to be even more extreme.

Second, while rational arguments can be made regarding whether re-classified or specific weighted options have more (or less) validity than crude reported numbers of deaths, the real gold standard validation of different estimates requires autopsy studies and in-depth audit of medical records. Error rates regarding causes of death in death certificates are so high that healthy skepticism is warranted. Illustratively, in Vermont, a medical record audit study found that the underlying cause of death reported in death certificates was wrong in 60% and errors in ICD codes appeared in 92% of certificates (33). There is substantial potential for errors even with detailed medical records. For example, a study of medical examiners (arguably, the best-trained group to certify deaths) in Australia found that when they attributed deaths based on detailed perusal of medical records, their selected cause of death was still wrong 28% of the time when compared to autopsy (34). Third, we provide separate results for two large age strata and these already show that the impact of different weighting may vary for some death causes in these age groups. However, we cannot exclude that the patterns generated from weighting may deviate even further from the baseline in more granular substrata, e.g. defined by residence status (community versus nursing homes) or socioeconomic indicators. Documentation of multiple as opposed to single causes of death depends on a wide range of socioeconomic and other factors (35). These associations may reflect both genuine differences in multi-morbidity, as well as differences in the likelihood that multiple causes would be documented even if they exist. For example, multiple causes may be more likely to be documented when patients get better health care with continuity and when they are hospitalized rather than when they have no care and die at home (35).

Acknowledging these sources of complexity, we argue that re-classification patterns and multiple causes of death should be routinely considered in analyses of death certificate data. Different re-classification patterns may point to structural defects in death certification that may require targeted training and education of certifiers; or that specific mortality coding rules may be promoting misleading changes. Statistics based on single underlying causes of death may be grossly inappropriate, more so for some causes like COVID-19 than for others. The reliance on single underlying causes of death has long served to encourage condition-specific focus in service provision, medical training, research and lobbying. Public health and health statistics worldwide should move away from unilateral dependence on this concept of only relative value and embrace more widely approaches that consider multiple causes of death. Different patterns emerging from weighting may point out that the burden from specific causes and categories of causes may be much larger or smaller than traditional death statistics suggest. This can have profound implications for resource allocation for both research and healthcare. Incorporation of weighted estimates may help shape more appropriate, evidence-based public health priorities. Additional in-depth validation assessments of death causes in diverse populations, including higher rates of autopsies, are also needed to better understand and more properly calibrate death certificate data.

## Data Availability

All data produced in the present study are available upon reasonable request to the authors.

https://github.com/scientific-computing-user/Reclassification-and-Weighting-of-Multiple-Causes-of-Death-US-Death-Certificates-2003-2023/

## SUPPLEMENTARY MATERIAL

### Supplementary Tables

**Table S1.** Causes of death classified in broad disease categories in Record Axis taken from. Table 1 **to show ratio of counts to Underlying.**

| Letter | Underlying | Contributing | Only | Anywhere | Contributing/<br>Underlying | Anywhere/<br>Underlying |
| --- | --- | --- | --- | --- | --- | --- |
| **All Ages** |  |  |  |  |  |  |
| **Circulatory** | 17,796,602 | 13,510,161 | 7,678,693 | 31,306,763 | 0.76 | 1.76 |
| **Cancer** | 12,576,466 | 1,435,404 | 5,482,941 | 14,011,870 | 0.11 | 1.11 |
| **Other Natural** | 10,708,481 | 18,124,458 | 4,662,437 | 28,832,939 | 1.69 | 2.69 |
| **Respiratory** | 5,303,862 | 9,896,044 | 1,295,750 | 15,199,906 | 1.87 | 2.87 |
| **Endocrine\*** | 2,560,585 | 6,780,460 | 156,081 | 9,341,045 | **2.65** | **3.65** |
| **Digestive** | 1,731,012 | 2,394,730 | 346,257 | 4,125,742 | 1.38 | 2.38 |
| **COVID-19** | 1,004,208 | 165,605 | 58,208 | 1,169,813 | 0.16 | 1.16 |
| **Drug Poisoning** | 1,229,766 | 241,136 | 701,531 | 1,470,902 | 0.20 | 1.20 |
| **Transport** | 915,055 | 19,491 | 853 | 934,546 | 0.02 | 1.02 |
| **Suicide** | 762,109 | 3,190 | 662 | 765,299 | 0.00 | 1.00 |
| **Alcohol-related** | 676,440 | 679,715 | 155,262 | 1,356,155 | 1.00 | 2.00 |
| **Other External** | 671,615 | 4,197,101 | 269,509 | 4,868,716 | **6.25** | **7.25** |
| **Falls** | 654,100 | 159,415 | 161 | 813,515 | 0.24 | 1.24 |
| **Homicide** | 396,530 | 4,118 | 275 | 400,648 | 0.01 | 1.01 |
| **Under 65** |  |  |  |  |  |  |
| **Circulatory** | 3,311,687 | 2,870,974 | 1,427,809 | 6,182,661 | 0.87 | 1.87 |
| **Cancer** | 3,634,924 | 217,427 | 1,706,537 | 3,852,351 | 0.06 | 1.06 |
| **Other Natural** | 2,085,650 | 4,228,619 | 1,017,129 | 6,314,269 | 2.03 | 3.03 |
| **Respiratory** | 808,320 | 2,139,295 | 175,255 | 2,947,615 | 2.65 | 3.65 |
| **Endocrine\*** | 721,155 | 1,548,828 | 66,023 | 2,269,983 | **2.15** | **3.15** |
| **Digestive** | 531,999 | 854,850 | 130,223 | 1,386,849 | 1.61 | 2.61 |
| **COVID-19** | 248,498 | 31,601 | 15,369 | 280,099 | 0.13 | 1.13 |
| **Drug Poisoning** | 1,159,365 | 205,731 | 673,252 | 1,365,096 | 0.18 | 1.18 |
| **Transport** | 744,515 | 10,074 | 704 | 754,589 | 0.01 | 1.01 |
| **Suicide** | 621,785 | 2,395 | 504 | 624,180 | 0.00 | 1.00 |
| **Alcohol-related** | 525,985 | 515,548 | 126,220 | 1,041,533 | 0.98 | 1.98 |
| **Other External** | 355,615 | 2,340,926 | 205,078 | 2,696,541 | **6.58** | **7.58** |
| **Falls** | 99,661 | 14,162 | 23 | 113,823 | 0.14 | 1.14 |
| **Homicide** | 376,112 | 2,709 | 256 | 378,821 | 0.01 | 1.01 |
| **65 Years or Older** |  |  |  |  |  |  |
| **Circulatory** | 14,484,915 | 10,639,187 | 6,250,884 | 25,124,102 | 0.73 | 1.73 |
| **Cancer** | 8,941,542 | 1,217,977 | 3,776,404 | 10,159,519 | 0.14 | 1.14 |
| **Other Natural** | 8,622,831 | 13,895,839 | 3,645,308 | 22,518,670 | 1.61 | 2.61 |
| **Respiratory** | 4,495,542 | 7,756,749 | 1,120,495 | 12,252,291 | 1.73 | 2.73 |
| **Endocrine\*** | 1,839,430 | 5,231,632 | 90,058 | 7,071,062 | **2.84** | **3.84** |
| **Digestive** | 1,199,013 | 1,539,880 | 216,034 | 2,738,893 | 1.28 | 2.28 |
| **COVID-19** | 755,710 | 134,004 | 42,839 | 889,714 | 0.18 | 1.18 |
| **Drug Poisoning** | 70,401 | 35,405 | 28,279 | 105,806 | 0.50 | 1.50 |
| **Transport** | 170,540 | 9,417 | 149 | 179,957 | 0.06 | 1.06 |
| **Suicide** | 140,324 | 795 | 158 | 141,119 | 0.01 | 1.01 |
| **Alcohol-related** | 150,455 | 164,167 | 29,042 | 314,622 | 1.09 | 2.09 |
| **Other External** | 316,000 | 1,856,175 | 64,431 | 2,172,175 | **5.87** | **6.87** |
| **Falls** | 554,439 | 145,253 | 138 | 699,692 | 0.26 | 1.26 |
| **Homicide** | 20,418 | 1,409 | 19 | 21,827 | 0.07 | 1.07 |

\*endocrine nutritional and metabolic

**Table S2.** Causes of death classified in broad disease categories in Record Axis and Entity Axis, USA 2003-2023 using the sensitivity analysis approach (choosing the first listed ICD code in the last line of the Part I for the Entity Axis)

| Disease | Record Axis |  |  |  | Entity Axis |  |  |  |
| --- | --- | --- | --- | --- | --- | --- | --- | --- |
| Disease | Underlying | Contributing | Only | Any | Underlying | Contributing | Only | Any |
| **Circulatory** | 17,796,602 (0%) | 13,510,161 | 7,678,693 | 31,306,763 | 17,770,736 | 13,605,199 | 7,521,247 | 31,375,935 |
| **Cancer** | 12,576,466 (12%) | 1,435,404 | 5,482,941 | 14,011,870 | 11,182,712 | 2,848,812 | 5,476,807 | 14,031,524 |
| **Other Natural** | 10,708,481 (-13%) | 18,124,458 | 4,662,437 | 28,832,939 | 12,342,764 | 17,215,274 | 4,603,744 | 29,558,038 |
| **Respiratory** | 5,303,862 (-11%) | 9,896,044 | 1,295,750 | 15,199,906 | 5,979,056 | 9,276,817 | 1,295,638 | 15,255,873 |
| **Endocrine\*** | 2,560,585 (19%) | 6,780,460 | 156,081 | 9,341,045 | 2,159,635 | 7,186,400 | 124,334 | 9,346,035 |
| **Digestive** | 1,731,012 (3%) | 2,394,730 | 346,257 | 4,125,742 | 1,674,443 | 2,693,506 | 346,111 | 4,367,949 |
| **COVID-19** | 1,004,208 (69%) | 165,605 | 58,208 | 1,169,813 | 595,609 | 574,204 | 58,211 | 1,169,813 |
| **Drug Poisoning** | 1,229,766 (5%) | 241,136 | 701,531 | 1,470,902 | 1,168,358 | 304,773 | 701,529 | 1,473,131 |
| **Transport** | 915,055 (430%) | 19,491 | 853 | 934,546 | 172,795 | 761,743 | 852 | 934,538 |
| **Suicide** | 762,109 (555%) | 3,190 | 662 | 765,299 | 116,440 | 648,857 | 662 | 765,297 |
| **Alcohol-related** | 676,440 (18%) | 679,715 | 155,262 | 1,356,155 | 571,405 | 784,750 | 102,929 | 1,356,155 |
| **Other External** | 671,615 (-78%) | 4,197,101 | 269,509 | 4,868,716 | 2,988,882 | 1,883,187 | 269,514 | 4,872,069 |
| **Falls** | 654,100 (178%) | 159,415 | 161 | 813,515 | 235,675 | 577,837 | 161 | 813,512 |
| **Homicide** | 396,530 (1300%) | 4,118 | 275 | 400,648 | 28,321 | 372,327 | 275 | 400,648 |
| **Uncertain** | 0 | 518 | 0 | 518 | 0 | 0 | 0 | 0 |
| **Total** | **56,986,831** | **57,611,546** | **20,808,620** | **114,598,377** | **56,986,831** | **58,733,686** | **20,502,014** | **115,720,517** |

\*endocrine nutritional and metabolic

**Table S3.** Changes in causes of death classified in broad disease categories in Entity Axis when using the first listed ICD code rather than the last, in the last line of Part I. There is no change on the Record Axis.

| **BROAD CATEGORY** | **Entity Axis**<br>**count using**<br>**first ICD-10** | **Entity Axis**<br>**count using**<br>**last ICD-10** | **Change**<br>**in count** | **Percent**<br>**Change**<br>**in count** |
| --- | --- | --- | --- | --- |
| **Circulatory** | 17,770,736 | 17,793,268 | -22,532 | 0% |
| **Cancer** | 11,182,712 | 11,214,012 | -31,300 | 0% |
| **Other Natural** | 12,342,764 | 12,386,337 | -43,573 | 0% |
| **Respiratory** | 5,979,056 | 5,978,426 | 630 | 0% |
| **Endocrine, Nutritional and Metabolic** | 2,159,635 | 2,209,113 | -49,478 | -2% |
| **Digestive** | 1,674,443 | 1,675,880 | -1,437 | 0% |
| **COVID-19** | 595,609 | 522,512 | 73,097 | 14% |
| **Drug Poisoning** | 1,168,358 | 1,130,962 | 37,396 | 3% |
| **Transport** | 172,795 | 637,778 | -464,983 | -73% |
| **Suicide** | 116,440 | 609,437 | -492,997 | -81% |
| **Alcohol-related** | 571,405 | 686,868 | -115,463 | -17% |
| **Other External** | 2,988,882 | 1,451,348 | 1,537,534 | 106% |
| **Falls** | 235,675 | 386,291 | -150,616 | -39% |
| **Homicide** | 28,321 | 304,599 | -276,278 | -91% |

**Table S4.** The 30 most frequent ICD-10 codes shown as underlying cause in the Entity Axis replaced with COVID-19 (ICD-10 code U071) as underlying cause in the Record Axis.

| ICD-10 code | Count | Cumulative percentage (%) | Description of cause |
| --- | --- | --- | --- |
| J189 | 220,367 | 43.8 | Pneumonia, unspecified |
| B99 | 16,742 | 47.2 | Other infectious diseases |
| A419 | 13,306 | 49.8 | Sepsis, unspecified organism |
| J449 | 12,556 | 52.3 | COPD, unspecified |
| J960 | 10,724 | 54.4 | Acute respiratory failure |
| N179 | 10,579 | 56.5 | Acute kidney failure |
| J969 | 10,470 | 58.6 | Respiratory failure, unspecified |
| I10 | 10,300 | 60.7 | Essential hypertension |
| F03 | 9,761 | 62.6 | Dementia, unspecified |
| J80 | 7,590 | 64.1 | ARDS |
| I500 | 6,199 | 65.4 | Congestive heart failure |
| J129 | 6,099 | 66.6 | Viral pneumonia, unspecified |
| I251 | 6,076 | 67.8 | Atherosclerotic heart disease |
| E149 | 5,587 | 68.9 | Diabetes, unspecified |
| B349 | 5,085 | 69.9 | Viral infection, unspecified |
| E119 | 4,267 | 70.8 | Type 2 diabetes |
| G309 | 4,245 | 71.6 | Alzheimer disease |
| I48 | 4,170 | 72.4 | Atrial fibrillation |
| R090 | 4,032 | 73.2 | Asphyxia |
| N288 | 3,907 | 74.0 | Other kidney disorders |
| I469 | 3,706 | 74.7 | Cardiac arrest |
| N189 | 3,538 | 75.4 | CKD, unspecified |
| I269 | 3,402 | 76.1 | Pulmonary embolism |
| R688 | 3,351 | 76.8 | Other specified symptoms |
| N185 | 3,076 | 77.4 | CKD stage 5 |
| I509 | 2,958 | 78.0 | Heart failure, unspecified |
| J159 | 2,924 | 78.6 | Bacterial pneumonia, unspecified |
| N19 | 2,575 | 79.1 | Renal failure, unspecified |
| J690 | 2,468 | 79.6 | Aspiration pneumonia |
| C349 | 2,112 | 80.0 | Lung cancer |

**Table S5A.** Changes between Record Axis and Entity Axis causes of death classified in broad disease categories when using the first listed ICD code in the last line of Part I of the Entity Axis versus using the last listed ICD code in the last line of Part I of the Entity Axis.

| Year | Record Axis count | Entity Axis ICD-10 count with first degenerate Entity Axis code |  | Entity Axis ICD-10 count with last degenerate Entity Axis code |  | Entity Axis ICD-10 count with first degenerate Entity Axis 3 character code |  | Entity Axis ICD-10 count with last degenerate Entity Axis 3 character code |  | Entity Axis broad categories count with first degenerate Entity Axis code |  | Entity Axis broad categories count with first degenerate Entity Axis code |  |
| --- | --- | --- | --- | --- | --- | --- | --- | --- | --- | --- | --- | --- | --- |
|  | in 1000s | in 1000s | Entity / Record % | in 1000s | Entity / Record % | in 1000s | Entity / Record | in 1000s | Entity / Record | in 1000s | Entity / Record | in 1000s | Entity / Record |
| 2003 | 2,448 | 1,759 | 71.8 | 1,733 | 70.8 | 1,781 | 72.8 | 1,755 | 71.7 | 2,131 | 87.0 | 2,078 | 84.9 |
| 2004 | 2,398 | 1,719 | 71.7 | 1,695 | 70.7 | 1,740 | 72.6 | 1,716 | 71.6 | 2,080 | 86.7 | 2,027 | 84.5 |
| 2005 | 2,448 | 1,762 | 72.0 | 1,737 | 71.0 | 1,784 | 72.9 | 1,759 | 71.8 | 2,123 | 86.7 | 2,067 | 84.5 |
| 2006 | 2,426 | 1,723 | 71.0 | 1,699 | 70.0 | 1,752 | 72.2 | 1,728 | 71.2 | 2,097 | 86.4 | 2,041 | 84.1 |
| 2007 | 2,424 | 1,726 | 71.2 | 1,703 | 70.3 | 1,749 | 72.1 | 1,725 | 71.2 | 2,090 | 86.2 | 2,031 | 83.8 |
| 2008 | 2,472 | 1,743 | 70.5 | 1,720 | 69.6 | 1,778 | 71.9 | 1,754 | 71.0 | 2,123 | 85.9 | 2,063 | 83.5 |
| 2009 | 2,437 | 1,721 | 70.6 | 1,698 | 69.7 | 1,755 | 72.0 | 1,732 | 71.0 | 2,094 | 85.9 | 2,035 | 83.5 |
| 2010 | 2,468 | 1,743 | 70.6 | 1,721 | 69.7 | 1,776 | 71.9 | 1,753 | 71.0 | 2,117 | 85.8 | 2,057 | 83.3 |
| 2011 | 2,515 | 1,765 | 70.2 | 1,742 | 69.3 | 1,805 | 71.7 | 1,781 | 70.8 | 2,155 | 85.7 | 2,094 | 83.2 |
| 2012 | 2,543 | 1,782 | 70.1 | 1,759 | 69.1 | 1,821 | 71.6 | 1,797 | 70.6 | 2,174 | 85.5 | 2,109 | 82.9 |
| 2013 | 2,597 | 1,816 | 69.9 | 1,791 | 69.0 | 1,860 | 71.6 | 1,834 | 70.6 | 2,221 | 85.5 | 2,155 | 83.0 |
| 2014 | 2,626 | 1,839 | 70.0 | 1,813 | 69.0 | 1,884 | 71.7 | 1,856 | 70.7 | 2,250 | 85.7 | 2,183 | 83.1 |
| 2015 | 2,713 | 1,888 | 69.6 | 1,860 | 68.6 | 1,934 | 71.3 | 1,904 | 70.2 | 2,319 | 85.5 | 2,247 | 82.8 |
| 2016 | 2,744 | 1,899 | 69.2 | 1,866 | 68.0 | 1,943 | 70.8 | 1,908 | 69.5 | 2,342 | 85.4 | 2,265 | 82.5 |
| 2017 | 2,814 | 1,921 | 68.3 | 1,885 | 67.0 | 1,968 | 70.0 | 1,930 | 68.6 | 2,386 | 84.8 | 2,309 | 82.1 |
| 2018 | 2,839 | 1,927 | 67.9 | 1,890 | 66.6 | 1,975 | 69.6 | 1,936 | 68.2 | 2,403 | 84.6 | 2,325 | 81.9 |
| 2019 | 2,855 | 1,933 | 67.7 | 1,893 | 66.3 | 1,981 | 69.4 | 1,939 | 67.9 | 2,418 | 84.7 | 2,337 | 81.9 |
| 2020 | 3,384 | 2,234 | 66.0 | 2,184 | 64.5 | 2,286 | 67.6 | 2,233 | 66.0 | 2,765 | 81.7 | 2,697 | 79.7 |
| 2021 | 3,464 | 2,203 | 63.6 | 2,149 | 62.0 | 2,253 | 65.0 | 2,196 | 63.4 | 2,747 | 79.3 | 2,694 | 77.8 |
| 2022 | 3,280 | 2,129 | 64.9 | 2,073 | 63.2 | 2,182 | 66.5 | 2,123 | 64.7 | 2,691 | 82.1 | 2,608 | 79.5 |
| 2023 | 3,091 | 2,031 | 65.7 | 1,977 | 64.0 | 2,083 | 67.4 | 2,026 | 65.6 | 2,586 | 83.7 | 2,495 | 80.7 |
| Total 2003-23 | 56,987 | 39,261 | 68.9 | 38,588 | 67.7 | 40,091 | 70.4 | 39,385 | 69.1 | 48,313 | 84.8 | 46,920 | 82.3 |
| Total 2003-19 | 43,768 | 30,663 | 70.1 | 30,205 | 69.0 | 31,287 | 71.5 | 30,807 | 70.4 | 37,523 | 85.7 | 36,425 | 83.2 |
| Total 2020-23 | 13,219 | 8,598 | 65.0 | 8,383 | 63.4 | 8,804 | 66.6 | 8,578 | 64.9 | 10,790 | 81.6 | 10,495 | 79.4 |
| Total 2003-23 | 56,986,831 | 39,260,709 | 68.9 | 38,588,288 | 67.7 | 40,090,579 | 70.4 | 39,384,860 | 69.1 | 48,313,403 | 84.8 | 46,919,850 | 82.3 |
| Total 2003-19 | 43,768,050 | 30,663,135 | 70.1 | 30,205,010 | 69.0 | 31,286,621 | 71.5 | 30,806,674 | 70.4 | 37,523,391 | 85.7 | 36,425,349 | 83.2 |
| Total 2020-23 | 13,218,781 | 8,597,574 | 65.0 | 8,383,278 | 63.4 | 8,803,958 | 66.6 | 8,578,186 | 64.9 | 10,790,012 | 81.6 | 10,494,501 | 79.4 |

**Table S5B.** Changes in causes of death classified in broad disease categories in Entity Axis when using the first listed ICD code in the last line of Part I versus using the last listed ICD code in the last line of Part I, showing results excluding COVID (U071).

| Year | Record<br>Axis count | Entity Axis ICD-10<br>count with first<br>degenerate Entity Axis<br>code |  | Entity Axis broad<br>categories count with<br>first degenerate Entity<br>Axis code |  | Entity Axis ICD-10<br>count with first<br>degenerate Entity Axis<br>code; excluding U071 |  | Entity Axis broad<br>categories count with first<br>degenerate Entity Axis<br>code; excluding U071 |  |
| --- | --- | --- | --- | --- | --- | --- | --- | --- | --- |
|  | in 1000s | in 1000s | Entity /<br>Record % | in 1000s | Entity /<br>Record % | in 1000s | Entity /<br>Record % | in 1000s | Entity /<br>Record % |
| 2003 | 2,448 | 1,759 | 71.8 | 2,131 | 87.0 | 1,759 | 71.8 | 2,131 | 87.0 |
| 2004 | 2,398 | 1,719 | 71.7 | 2,080 | 86.7 | 1,719 | 71.7 | 2,080 | 86.7 |
| 2005 | 2,448 | 1,762 | 72.0 | 2,123 | 86.7 | 1,762 | 72.0 | 2,123 | 86.7 |
| 2006 | 2,426 | 1,723 | 71.0 | 2,097 | 86.4 | 1,723 | 71.0 | 2,097 | 86.4 |
| 2007 | 2,424 | 1,726 | 71.2 | 2,090 | 86.2 | 1,726 | 71.2 | 2,090 | 86.2 |
| 2008 | 2,472 | 1,743 | 70.5 | 2,123 | 85.9 | 1,743 | 70.5 | 2,123 | 85.9 |
| 2009 | 2,437 | 1,721 | 70.6 | 2,094 | 85.9 | 1,721 | 70.6 | 2,094 | 85.9 |
| 2010 | 2,468 | 1,743 | 70.6 | 2,117 | 85.8 | 1,743 | 70.6 | 2,117 | 85.8 |
| 2011 | 2,515 | 1,765 | 70.2 | 2,155 | 85.7 | 1,765 | 70.2 | 2,155 | 85.7 |
| 2012 | 2,543 | 1,782 | 70.1 | 2,174 | 85.5 | 1,782 | 70.1 | 2,174 | 85.5 |
| 2013 | 2,597 | 1,816 | 69.9 | 2,221 | 85.5 | 1,816 | 69.9 | 2,221 | 85.5 |
| 2014 | 2,626 | 1,839 | 70.0 | 2,250 | 85.7 | 1,839 | 70.0 | 2,250 | 85.7 |
| 2015 | 2,713 | 1,888 | 69.6 | 2,319 | 85.5 | 1,888 | 69.6 | 2,319 | 85.5 |
| 2016 | 2,744 | 1,899 | 69.2 | 2,342 | 85.4 | 1,899 | 69.2 | 2,342 | 85.4 |
| 2017 | 2,814 | 1,921 | 68.3 | 2,386 | 84.8 | 1,921 | 68.3 | 2,386 | 84.8 |
| 2018 | 2,839 | 1,927 | 67.9 | 2,403 | 84.6 | 1,927 | 67.9 | 2,403 | 84.6 |
| 2019 | 2,855 | 1,933 | 67.7 | 2,418 | 84.7 | 1,933 | 67.7 | 2,418 | 84.7 |
| 2020 | 3,384 | 2,234 | 66.0 | 2,765 | 81.7 | 2,026 | 66.9 | 2,556 | 84.5 |
| 2021 | 3,464 | 2,203 | 63.6 | 2,747 | 79.3 | 2,022 | 66.4 | 2,566 | 84.3 |
| 2022 | 3,280 | 2,129 | 64.9 | 2,691 | 82.1 | 2,041 | 66.1 | 2,603 | 84.3 |
| 2023 | 3,091 | 2,031 | 65.7 | 2,586 | 83.7 | 2,008 | 66.1 | 2,563 | 84.4 |
| Total 2003-23 | 56,987 | 39,261 | 68.9 | 48,313 | 84.8 | 38,759 | 69.3 | 47,812 | 85.4 |
| Total 2003-19 | 43,768 | 30,663 | 70.1 | 37,523 | 85.7 | 30,663 | 70.1 | 37,523 | 85.7 |
| Total 2020-23 | 13,219 | 8,598 | 65.0 | 10,790 | 81.6 | 8,096 | 66.4 | 10,288 | 84.4 |
| Total 2003-23 | 56,986,831 | 39,260,709 | 68.9 | 48,313,403 | 84.8 | 38,759,268 | 69.3 | 47,811,656 | 85.4 |
| Total 2003-19 | 43,768,050 | 30,663,135 | 70.1 | 37,523,391 | 85.7 | 30,663,135 | 70.1 | 37,523,391 | 85.7 |
| Total 2020-23 | 13,218,781 | 8,597,574 | 65.0 | 10,790,012 | 81.6 | 8,096,133 | 66.4 | 10,288,265 | 84.4 |

**Table S6.** Top five ICD-10 codes boosting the counts for Other External causes of death when contributing causes are included by weighting.

| Count | Cumulative % | ICD-10 | Explanation | Count | Cumulative % | ICD-10 | Explanation |
| --- | --- | --- | --- | --- | --- | --- | --- |
| **Under 65 years. Other External is not underlying cause of death** |  |  |  | **Under 65 years. Other External is underlying cause of death** |  |  |  |
| 41026 | 3.5% | S099 | Unspecified injury of head | 4656 | 2.6% | R568 | Other and unspecified convulsions |
| 38155 | 6.8% | T179 | Foreign body in respiratory tract, part unspecified | 4033 | 4.8% | S099 | Unspecified injury of head |
| 31183 | 9.5% | T509 | Poisoning: Other and unspecified drugs, medicaments and biological substances | 2881 | 6.4% | I469 | Cardiac arrest, unspecified |
| 18045 | 11.0% | S069 | Intracranial injury, unspecified | 2768 | 7.9% | T68 | Hypothermia |
| 13391 | 12.1% | I469 | Cardiac arrest, unspecified | 2638 | 9.4% | T510 | Toxic effect: Ethanol |
| **65 years or over. Other External is not underlying cause of death** |  |  |  | **65 years or over. Other External is underlying cause of death** |  |  |  |
| 125171 | 15.4% | T179 | Foreign body in respiratory tract, part unspecified | 7431 | 0.4% | J969 | Respiratory failure, unspecified |
| 67061 | 19.1% | S720 | Fracture of neck of femur | 6138 | 0.7% | R628 | Other lack of expected normal physiological development |
| 40565 | 21.3% | J969 | Respiratory failure, unspecified | 5065 | 1.0% | J449 | Chronic obstructive pulmonary disease, unspecified |
| 39975 | 23.5% | J449 | Chronic obstructive pulmonary disease, unspecified | 4877 | 1.3% | R13 | Dysphagia |
| 33167 | 25.3% | R13 | Dysphagia | 4433 | 1.5% | R688 | Other specified general symptoms and signs |

**Table S7.**
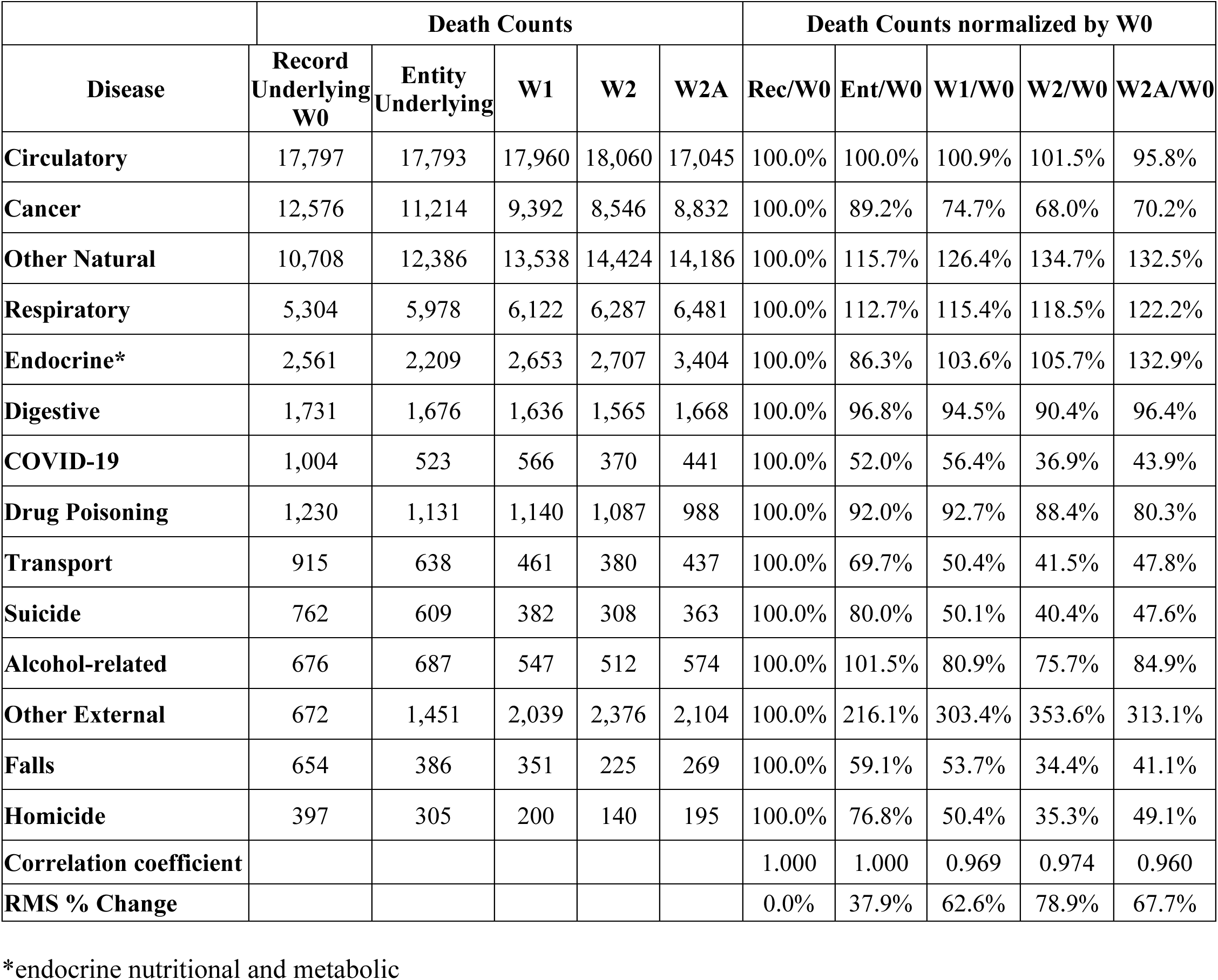
Comparing the percentage changes in death counts between the Entity and Record axes with those between unweighted and weighted counts.

**Table S8.** Slopes of monthly death counts for different diseases, age buckets and weighting schemes. We only include those instances where the R2 for both the linear and logarithmic fit is greater than 0.6.

| Disease | Bucket | Scheme | R2 | B<br>linear /<br>y % | B log / y<br>% | Disease | Bucket | Scheme | R2 | B linear<br>/ y % | B log / y<br>% |
| --- | --- | --- | --- | --- | --- | --- | --- | --- | --- | --- | --- |
| Circulatory | GE65 | W0 | 0.694 | -2.5 | -2.4 | Drug poisoning | LT65 | W2A | 0.833 | 6.8 | 6.8 |
| Circulatory | GE65 | W1 | 0.637 | -2.1 | -2.0 | Suicide | ALL | W0 | 0.771 | 2.0 | 2.1 |
| Circulatory | GE65 | W2A | 0.606 | -2.0 | -1.9 | Suicide | ALL | W1 | 0.765 | 2.0 | 2.0 |
| Cancer | ALL | W2 | 0.637 | -0.7 | -0.7 | Suicide | ALL | W2 | 0.678 | 1.7 | 1.7 |
| Cancer | ALL | W2A | 0.646 | -0.8 | -0.7 | Suicide | ALL | W2A | 0.769 | 2.0 | 2.1 |
| Cancer | GE65 | W0 | 0.908 | -1.9 | -1.9 | Suicide | LT65 | W0 | 0.786 | 2.1 | 2.2 |
| Cancer | GE65 | W1 | 0.924 | -2.1 | -2.0 | Suicide | LT65 | W1 | 0.782 | 2.1 | 2.2 |
| Cancer | GE65 | W2 | 0.938 | -2.3 | -2.3 | Suicide | LT65 | W2 | 0.710 | 1.9 | 1.9 |
| Cancer | GE65 | W2A | 0.938 | -2.3 | -2.3 | Suicide | LT65 | W2A | 0.784 | 2.1 | 2.1 |
| Cancer | LT65 | W0 | 0.602 | -0.9 | -0.9 | Alcohol-related | ALL | W0 | 0.856 | 4.7 | 4.7 |
| Cancer | LT65 | W1 | 0.762 | -1.2 | -1.2 | Alcohol-related | ALL | W1 | 0.855 | 4.5 | 4.4 |
| Cancer | LT65 | W2 | 0.827 | -1.5 | -1.6 | Alcohol-related | ALL | W2 | 0.849 | 4.2 | 4.2 |
| Cancer | LT65 | W2A | 0.826 | -1.5 | -1.6 | Alcohol-related | ALL | W2A | 0.844 | 4.4 | 4.3 |
| Other natural | ALL | W0 | 0.629 | 1.9 | 1.9 | Alcohol-related | GE65 | W0 | 0.818 | 4.3 | 4.2 |
| Other natural | ALL | W1 | 0.617 | 1.8 | 1.9 | Alcohol-related | GE65 | W1 | 0.824 | 4.2 | 4.1 |
| Other natural | ALL | W2 | 0.618 | 1.9 | 1.9 | Alcohol-related | GE65 | W2 | 0.816 | 4.0 | 3.9 |
| Endocrine | LT65 | W0 | 0.642 | 2.5 | 2.4 | Alcohol-related | GE65 | W2A | 0.811 | 4.2 | 4.1 |
| Endocrine | LT65 | W1 | 0.625 | 3.0 | 2.9 | Alcohol-related | LT65 | W0 | 0.846 | 4.5 | 4.5 |
| Endocrine | LT65 | W2 | 0.609 | 3.3 | 3.1 | Alcohol-related | LT65 | W1 | 0.844 | 4.3 | 4.3 |
| Endocrine | LT65 | W2A | 0.618 | 3.1 | 3.0 | Alcohol-related | LT65 | W2 | 0.838 | 4.1 | 4.1 |
| Drug poisoning | ALL | W0 | 0.844 | 6.7 | 6.7 | Alcohol-related | LT65 | W2A | 0.835 | 4.2 | 4.2 |
| Drug poisoning | ALL | W1 | 0.84 | 6.7 | 6.7 | Falls | ALL | W0 | 0.952 | 4.3 | 4.5 |
| Drug poisoning | ALL | W2 | 0.837 | 6.7 | 6.7 | Falls | ALL | W1 | 0.947 | 4.2 | 4.4 |
| Drug poisoning | ALL | W2A | 0.835 | 6.7 | 6.6 | Falls | ALL | W2 | 0.940 | 3.7 | 3.8 |
| Drug poisoning | GE65 | W0 | 0.818 | 8.3 | 8.3 | Falls | ALL | W2A | 0.938 | 3.8 | 3.9 |
| Drug poisoning | GE65 | W1 | 0.818 | 8.5 | 8.6 | Falls | GE65 | W0 | 0.854 | 2.8 | 3.0 |
| Drug poisoning | GE65 | W2 | 0.816 | 8.7 | 8.7 | Falls | GE65 | W1 | 0.852 | 2.7 | 2.8 |
| Drug poisoning | GE65 | W2A | 0.82 | 8.8 | 8.8 | Falls | GE65 | W2 | 0.773 | 2.0 | 2.1 |
| Drug poisoning | LT65 | W0 | 0.842 | 6.8 | 6.9 | Falls | GE65 | W2A | 0.802 | 2.2 | 2.3 |
| Drug poisoning | LT65 | W1 | 0.838 | 6.9 | 6.9 | Falls | LT65 | W0 | 0.666 | 1.9 | 1.9 |
| Drug poisoning | LT65 | W2 | 0.835 | 6.9 | 6.8 | Falls | LT65 | W1 | 0.680 | 1.9 | 1.9 |

\*endocrine nutritional and metabolic

**Table S9.** Mapping of ICD-10 codes to Broad disease categories as adapted from Wrigley-Field (Wrigley-Field E, Raquib RV, Berry KM, Morris KJ, Stokes AC. Mortality trends among early adults in the United States, 1999-2023. JAMA Network Open 2025:8e2457538).

| ICD-10 | Broad disease category | First_letter | Description |
| --- | --- | --- | --- |
| I169 | Circulatory | I | I16.9 (Hypertensive crisis, unspecified) |
| K85 | Digestive | K | K85 (Acute pancreatitis) |
| U099 | COVID-19 | U | U09.9 (Post COVID-19 condition) |
| E244 | Alcohol-related | A | E24.4 (Alcohol-induced pseudo-Cushing syndrome) |
| F10 | Alcohol-related | A | F10 (Mental and behavioural disorders due to use of alcohol) |
| G312 | Alcohol-related | A | G31.2 (Degeneration of nervous system due to alcohol) |
| G621 | Alcohol-related | A | G62.1 (Alcoholic polyneuropathy) |
| G721 | Alcohol-related | A | G72.1 (Alcoholic myopathy) |
| I426 | Alcohol-related | A | I42.6 (Alcoholic cardiomyopathy) |
| K292 | Alcohol-related | A | K29.2 (Alcoholic gastritis) |
| K70 | Alcohol-related | A | K70 (Alcoholic liver disease) |
| K852 | Alcohol-related | A | K85.2 (Alcohol-induced acute pancreatitis) |
| K860 | Alcohol-related | A | K86.0 (Alcohol-induced chronic pancreatitis) |
| R780 | Alcohol-related | A | R78.0 (Finding of alcohol in blood) |
| X45 | Alcohol-related | A | X45 (Accidental poisoning by and exposure to alcohol) |
| X65 | Alcohol-related | A | X65 (Intentional self-poisoning by and exposure to alcohol) |
| Y15 | Alcohol-related | A | Y15 (Poisoning by and exposure to alcohol, undetermined intent) |
| C00-D48 | Cancer | C | C00-D48 (Neoplasms) |
| U071 | COVID-19 | C | U07.1 (COVID-19) |
| K00-K14 | Digestive | D | K00-K14 (Diseases of oral cavity, salivary glands and jaws) |
| K20 | Digestive | D | K20 (Oesophagitis) |
| K21 | Digestive | D | K21 (Gastro-oesophageal reflux disease) |
| K22 | Digestive | D | K22 (Other diseases of oesophagus) |
| K25 | Digestive | D | K25 (Gastric ulcer) |
| K26 | Digestive | D | K26 (Duodenal ulcer) |
| K27 | Digestive | D | K27 (Peptic ulcer, site unspecified) |
| K28 | Digestive | D | K28 (Gastrojejunal ulcer) |
| K290 | Digestive | D | K29.0 (Acute haemorrhagic gastritis) |
| K291 | Digestive | D | K29.1 (Other acute gastritis) |
| K293 | Digestive | D | K29.3 (Chronic superficial gastritis) |
| K294 | Digestive | D | K29.4 (Chronic atrophic gastritis) |
| K295 | Digestive | D | K29.5 (Chronic gastritis, unspecified) |
| K296 | Digestive | D | K29.6 (Other gastritis) |
| K297 | Digestive | D | K29.7 (Gastritis, unspecified) |
| K298 | Digestive | D | K29.8 (Duodenitis) |
| K299 | Digestive | D | K29.9 (Gastroduodenitis, unspecified) |
| K30 | Digestive | D | K30 (Dyspepsia) |
| K31 | Digestive | D | K31 (Other diseases of stomach and duodenum) |
| K35-K38 | Digestive | D | K35-K38 (Diseases of appendix) |
| K40-K46 | Digestive | D | K40-K46 (Hernia) |
| K50-K52 | Digestive | D | K50-K52 (Noninfective enteritis and colitis) |
| K55-K63 | Digestive | D | K55-K63 (Other diseases of intestines) |
| K65-K66 | Digestive | D | K65-K66 (Diseases of peritoneum) |
| K71 | Digestive | D | K71 (Toxic liver disease) |
| K72 | Digestive | D | K72 (Hepatic failure, not elsewhere classified) |
| K73 | Digestive | D | K73 (Chronic hepatitis, not elsewhere classified) |
| K74 | Digestive | D | K74 (Fibrosis and cirrhosis of liver) |
| K75 | Digestive | D | K75 (Other inflammatory liver diseases) |
| K76 | Digestive | D | K76 (Other diseases of liver) |
| K80 | Digestive | D | K80 (Cholelithiasis) |
| **K81** | Digestive | D | K81 (Cholecystitis) |
| **K82** | Digestive | D | K82 (Other diseases of gallbladder) |
| **K83** | Digestive | D | K83 (Other diseases of biliary tract) |
| **K850** | Digestive | D | K85.0 (Idiopathic acute pancreatitis) |
| **K851** | Digestive | D | K85.1 (Biliary acute pancreatitis) |
| **K853** | Digestive | D | K85.3 (Drug-induced acute pancreatitis) |
| **K858** | Digestive | D | K85.8 (Other acute pancreatitis) |
| **K859** | Digestive | D | K85.9 (Acute pancreatitis, unspecified) |
| **K861** | Digestive | D | K86.1 (Other chronic pancreatitis) |
| **K862** | Digestive | D | K86.2 (Cyst of pancreas) |
| **K863** | Digestive | D | K86.3 (Pseudocyst of pancreas) |
| **K868** | Digestive | D | K86.8 (Other specified diseases of pancreas) |
| **K869** | Digestive | D | K86.9 (Disease of pancreas, unspecified) |
| **K90-K92** | Digestive | D | K90-K92 (Other diseases of the digestive system) |
| **F11** | Drug poisoning | D | F11 (Mental and behavioural disorders due to use of opioids) |
| **F12** | Drug poisoning | D | F12 (Mental and behavioural disorders due to use of cannabinoids) |
| **F13** | Drug poisoning | D | F13 (Mental and behavioural disorders due to use of sedatives or hypnotics) |
| **F14** | Drug poisoning | D | F14 (Mental and behavioural disorders due to use of cocaine) |
| **F15** | Drug poisoning | D | F15 (Mental and behavioural disorders due to use of other stimulants, including caffeine) |
| **F16** | Drug poisoning | D | F16 (Mental and behavioural disorders due to use of hallucinogens) |
| **F19** | Drug poisoning | D | F19 (Mental and behavioural disorders due to multiple drug use and use of other psychoactive substances) |
| **X60** | Drug poisoning | D | X60 (Intentional self-poisoning by and exposure to nonopioid analgesics, antipyretics and antirheumatics) |
| **X61** | Drug poisoning | D | X61 (Intentional self-poisoning by and exposure to antiepileptic, sedative-hypnotic, antiparkinsonism and psychotropic drugs, not elsewhere classified) |
| **X62** | Drug poisoning | D | X62 (Intentional self-poisoning by and exposure to narcotics and psychodysleptics [hallucinogens], not elsewhere classified) |
| **X63** | Drug poisoning | D | X63 (Intentional self-poisoning by and exposure to other drugs acting on the autonomic nervous system) |
| **X64** | Drug poisoning | D | X64 (Intentional self-poisoning by and exposure to other and unspecified drugs, medicaments and biological substances) |
| **X85** | Drug poisoning | D | X85 (Assault by drugs, medicaments and biological substances) |
| **Y10** | Drug poisoning | D | Y10 (Poisoning by and exposure to nonopioid analgesics, antipyretics and antirheumatics, undetermined intent) |
| **Y11** | Drug poisoning | D | Y11 (Poisoning by and exposure to antiepileptic, sedative-hypnotic, antiparkinsonism and psychotropic drugs, not elsewhere classified, undetermined intent) |
| **Y12** | Drug poisoning | D | Y12 (Poisoning by and exposure to narcotics and psychodysleptics [hallucinogens], not elsewhere classified, undetermined intent) |
| **Y13** | Drug poisoning | D | Y13 (Poisoning by and exposure to other drugs acting on the autonomic nervous system, undetermined intent) |
| **Y14** | Drug poisoning | D | Y14 (Poisoning by and exposure to other and unspecified drugs, medicaments and biological substances, undetermined intent) |
| **X40** | Drug poisoning | D | X40 (Accidental poisoning by and exposure to nonopioid analgesics, antipyretics and antirheumatics) |
| **X41** | Drug poisoning | D | X41 (Accidental poisoning by and exposure to antiepileptic, sedative-hypnotic, antiparkinsonism and psychotropic drugs, not elsewhere classified) |
| **X42** | Drug poisoning | D | X42 (Accidental poisoning by and exposure to narcotics and psychodysleptics [hallucinogens], not elsewhere classified) |
| **X43** | Drug poisoning | D | X43 (Accidental poisoning by and exposure to other drugs acting on the autonomic nervous system) |
| **X44** | Drug poisoning | D | X44 (Accidental poisoning by and exposure to other and unspecified drugs, medicaments and biological substances) |
| **E00-E07** | Endocrine | E | E00-E07 (Disorders of thyroid gland) |
| **E10-E14** | Endocrine | E | E10-E14 (Diabetes mellitus) |
| **E15-E16** | Endocrine | E | E15-E16 (Other disorders of glucose regulation and pancreatic internal secretion) |
| **E20** | Endocrine | E | E20 (Hypoparathyroidism) |
| **E21** | Endocrine | E | E21 (Hyperparathyroidism and other disorders of parathyroid gland) |
| **E22** | Endocrine | E | E22 (Hyperfunction of pituitary gland) |
| **E23** | Endocrine | E | E23 (Hypofunction and other disorders of pituitary gland) |
| **E240** | Endocrine | E | E24.0 (Pituitary-dependent Cushing disease) |
| **E241** | Endocrine | E | E24.1 (Nelson syndrome) |
| **E242** | Endocrine | E | E24.2 (Drug-induced Cushing syndrome) |
| **E243** | Endocrine | E | E24.3 (Ectopic ACTH syndrome) |
| **E248** | Endocrine | E | E24.8 (Other Cushing syndrome) |
| **E249** | Endocrine | E | E24.9 (Cushing syndrome, unspecified) |
| **E25** | Endocrine | E | E25 (Adrenogenital disorders) |
| **E26** | Endocrine | E | E26 (Hyperaldosteronism) |
| **E27** | Endocrine | E | E27 (Other disorders of adrenal gland) |
| **E28** | Endocrine | E | E28 (Ovarian dysfunction) |
| **E29** | Endocrine | E | E29 (Testicular dysfunction) |
| **E30** | Endocrine | E | E30 (Disorders of puberty, not elsewhere classified) |
| **E31** | Endocrine | E | E31 (Polyglandular dysfunction) |
| **E32** | Endocrine | E | E32 (Diseases of thymus) |
| **E34** | Endocrine | E | E34 (Other endocrine disorders) |
| **E40-E46** | Endocrine | E | E40-E46 (Malnutrition) |
| **E50-E64** | Endocrine | E | E50-E64 (Other nutritional deficiencies) |
| **E65-E68** | Endocrine | E | E65-E68 (Obesity and other hyperalimentation) |
| **E70-E88** | Endocrine | E | E70-E88 (Metabolic disorders) |
| **X86** | Homicide | H | X86 (Assault by corrosive substance) |
| **X87** | Homicide | H | X87 (Assault by pesticides) |
| **X88** | Homicide | H | X88 (Assault by gases and vapours) |
| **X89** | Homicide | H | X89 (Assault by other specified chemicals and noxious substances) |
| **X90** | Homicide | H | X90 (Assault by unspecified chemical or noxious substance) |
| **X91** | Homicide | H | X91 (Assault by hanging, strangulation and suffocation) |
| **X92** | Homicide | H | X92 (Assault by drowning and submersion) |
| **X93** | Homicide | H | X93 (Assault by handgun discharge) |
| **X94** | Homicide | H | X94 (Assault by rifle, shotgun and larger firearm discharge) |
| **X95** | Homicide | H | X95 (Assault by other and unspecified firearm discharge) |
| **X96** | Homicide | H | X96 (Assault by explosive material) |
| **X97** | Homicide | H | X97 (Assault by smoke, fire and flames) |
| **X98** | Homicide | H | X98 (Assault by steam, hot vapours and hot objects) |
| **X99** | Homicide | H | X99 (Assault by sharp object) |
| **Y00** | Homicide | H | Y00 (Assault by blunt object) |
| **Y01** | Homicide | H | Y01 (Assault by pushing from high place) |
| **Y02** | Homicide | H | Y02 (Assault by pushing or placing victim before moving object) |
| **Y03** | Homicide | H | Y03 (Assault by crashing of motor vehicle) |
| **Y04** | Homicide | H | Y04 (Assault by bodily force) |
| **Y05** | Homicide | H | Y05 (Sexual assault by bodily force) |
| **Y06** | Homicide | H | Y06 (Neglect and abandonment) |
| **Y07** | Homicide | H | Y07 (Other maltreatment syndromes) |
| **Y08** | Homicide | H | Y08 (Assault by other specified means) |
| **Y09** | Homicide | H | Y09 (Assault by unspecified means) |
| **Y871** | Homicide | H | Y87.1 (Sequelae of assault) |
| **I10-I15** | Circulatory | C | I10-I15 (Hypertensive diseases) |
| **I00-I02** | Circulatory | C | I00-I02 (Acute rheumatic fever) |
| **I05-I09** | Circulatory | C | I05-I09 (Chronic rheumatic heart diseases) |
| **I20-I25** | Circulatory | C | I20-I25 (Ischaemic heart diseases) |
| **I26-I28** | Circulatory | C | I26-I28 (Pulmonary heart disease and diseases of pulmonary circulation) |
| **I30** | Circulatory | C | I30 (Acute pericarditis) |
| **I31** | Circulatory | C | I31 (Other diseases of pericardium) |
| **I33** | Circulatory | C | I33 (Acute and subacute endocarditis) |
| I34 | Circulatory | C | I34 (Nonrheumatic mitral valve disorders) |
| I35 | Circulatory | C | I35 (Nonrheumatic aortic valve disorders) |
| I36 | Circulatory | C | I36 (Nonrheumatic tricuspid valve disorders) |
| I37 | Circulatory | C | I37 (Pulmonary valve disorders) |
| I38 | Circulatory | C | I38 (Endocarditis, valve unspecified) |
| I40 | Circulatory | C | I40 (Acute myocarditis) |
| I420 | Circulatory | C | I42.0 (Dilated cardiomyopathy) |
| I421 | Circulatory | C | I42.1 (Obstructive hypertrophic cardiomyopathy) |
| I422 | Circulatory | C | I42.2 (Other hypertrophic cardiomyopathy) |
| I423 | Circulatory | C | I42.3 (Endomyocardial (eosinophilic) disease) |
| I424 | Circulatory | C | I42.4 (Endocardial fibroelastosis) |
| I425 | Circulatory | C | I42.5 (Other restrictive cardiomyopathy) |
| I427 | Circulatory | C | I42.7 (Cardiomyopathy due to drugs and other external agents) |
| I428 | Circulatory | C | I42.8 (Other cardiomyopathies) |
| I429 | Circulatory | C | I42.9 (Cardiomyopathy, unspecified) |
| I44 | Circulatory | C | I44 (Atrioventricular and left bundle-branch block) |
| I45 | Circulatory | C | I45 (Other conduction disorders) |
| I46 | Circulatory | C | I46 (Cardiac arrest) |
| I47 | Circulatory | C | I47 (Paroxysmal tachycardia) |
| I48 | Circulatory | C | I48 (Atrial fibrillation and flutter) |
| I49 | Circulatory | C | I49 (Other cardiac arrhythmias) |
| I50 | Circulatory | C | I50 (Heart failure) |
| I51 | Circulatory | C | I51 (Complications and ill-defined descriptions of heart disease) |
| I60-I69 | Circulatory | C | I60-I69 (Cerebrovascular diseases) |
| I70-I78 | Circulatory | C | I70-I78 (Diseases of arteries, arterioles and capillaries) |
| I80-I89 | Circulatory | C | I80-I89 (Diseases of veins, lymphatic vessels and lymph nodes, not elsewhere classified) |
| I95-I99 | Circulatory | C | I95-I99 (Other and unspecified disorders of the circulatory system) |
| B20-B24 | Other natural | O | B20-B24 (Human immunodeficiency virus [HIV] disease) |
| F01-F09 | Other natural | O | F01-F09 (Organic, including symptomatic, mental disorders) |
| F17 | Other natural | O | F17 (Mental and behavioural disorders due to use of tobacco) |
| F18 | Other natural | O | F18 (Mental and behavioural disorders due to use of volatile solvents) |
| F20-F29 | Other natural | O | F20-F29 (Schizophrenia, schizotypal and delusional disorders) |
| F30-F39 | Other natural | O | F30-F39 (Mood [affective] disorders) |
| F40-F48 | Other natural | O | F40-F48 (Neurotic, stress-related and somatoform disorders) |
| F50-F59 | Other natural | O | F50-F59 (Behavioural syndromes associated with physiological disturbances and physical factors) |
| F60-F69 | Other natural | O | F60-F69 (Disorders of adult personality and behaviour) |
| F70-F79 | Other natural | O | F70-F79 (Mental retardation) |
| F80-F89 | Other natural | O | F80-F89 (Disorders of psychological development) |
| F90-F98 | Other natural | O | F90-F98 (Behavioural and emotional disorders with onset usually occurring in childhood and adolescence) |
| F99-F99 | Other natural | O | F99-F99 (Unspecified mental disorder) |
| A00-A09 | Other natural | O | A00-A09 (Intestinal infectious diseases) |
| A16-A19 | Other natural | O | A16-A19 (Tuberculosis) |
| A20-A28 | Other natural | O | A20-A28 (Certain zoonotic bacterial diseases) |
| A30-A49 | Other natural | O | A30-A49 (Other bacterial diseases) |
| A50-A64 | Other natural | O | A50-A64 (Infections with a predominantly sexual mode of transmission) |
| A65-A69 | Other natural | O | A65-A69 (Other spirochaetal diseases) |
| A70-A74 | Other natural | O | A70-A74 (Other diseases caused by chlamydiae) |
| A75-A79 | Other natural | O | A75-A79 (Rickettsioses) |
| A80-A89 | Other natural | O | A80-A89 (Viral infections of the central nervous system) |
| A90-A99 | Other natural | O | A90-A99 (Arthropod-borne viral fevers and viral haemorrhagic fevers) |
| B00-B09 | Other natural | O | B00-B09 (Viral infections characterized by skin and mucous membrane lesions) |
| B15-B19 | Other natural | O | B15-B19 (Viral hepatitis) |
| B25-B34 | Other natural | O | B25-B34 (Other viral diseases) |
| **B35-B49** | Other natural | O | B35-B49 (Mycoses) |
| **B50-B64** | Other natural | O | B50-B64 (Protozoal diseases) |
| **B65-B83** | Other natural | O | B65-B83 (Helminthiasis) |
| **B85-B89** | Other natural | O | B85-B89 (Pediculosis, acariasis and other infestations) |
| **B90-B94** | Other natural | O | B90-B94 (Sequelae of infectious and parasitic diseases) |
| **B99-B99** | Other natural | O | B99-B99 (Other infectious diseases) |
| **D50-D89** | Other natural | O | D50-D89 (Diseases of the blood and blood-forming organs and certain disorders involving the immune mechanism) |
| **G00-G09** | Other natural | O | G00-G09 (Inflammatory diseases of the central nervous system) |
| **G10-G14** | Other natural | O | G10-G14 (Systemic atrophies primarily affecting the central nervous system) |
| **G20-G25** | Other natural | O | G20-G25 (Extrapyramidal and movement disorders) |
| **G30** | Other natural | O | G30 (Alzheimer disease) |
| **G310** | Other natural | O | G31.0 (Circumscribed brain atrophy) |
| **G311** | Other natural | O | G31.1 (Senile degeneration of brain, not elsewhere classified) |
| **G318** | Other natural | O | G31.8 (Other specified degenerative diseases of nervous system) |
| **G319** | Other natural | O | G31.9 (Degenerative disease of nervous system, unspecified) |
| **G35-G37** | Other natural | O | G35-G37 (Demyelinating diseases of the central nervous system) |
| **G40-G47** | Other natural | O | G40-G47 (Episodic and paroxysmal disorders) |
| **G50-G58** | Other natural | O | G50-G58 (Nerve, nerve root and plexus disorders) |
| **G60** | Other natural | O | G60 (Hereditary and idiopathic neuropathy) |
| **G61** | Other natural | O | G61 (Inflammatory polyneuropathy) |
| **G620** | Other natural | O | G62.0 (Drug-induced polyneuropathy) |
| **G622** | Other natural | O | G62.2 (Polyneuropathy due to other toxic agents) |
| **G628** | Other natural | O | G62.8 (Other specified polyneuropathies) |
| **G629** | Other natural | O | G62.9 (Polyneuropathy, unspecified) |
| **G64** | Other natural | O | G64 (Other disorders of peripheral nervous system) |
| **G70** | Other natural | O | G70 (Myasthenia gravis and other myoneural disorders) |
| **G71** | Other natural | O | G71 (Primary disorders of muscles) |
| **G720** | Other natural | O | G72.0 (Drug-induced myopathy) |
| **G722** | Other natural | O | G72.2 (Myopathy due to other toxic agents) |
| **G723** | Other natural | O | G72.3 (Periodic paralysis) |
| **G724** | Other natural | O | G72.4 (Inflammatory myopathy, not elsewhere classified) |
| **G728** | Other natural | O | G72.8 (Other specified myopathies) |
| **G729** | Other natural | O | G72.9 (Myopathy, unspecified) |
| **G80-G83** | Other natural | O | G80-G83 (Cerebral palsy and other paralytic syndromes) |
| **G90-G98** | Other natural | O | G90-G98 (Other disorders of the nervous system) |
| **H00-H57** | Other natural | O | H00-H57 (Diseases of the eye and adnexa) |
| **H60-H93** | Other natural | O | H60-H93 (Diseases of the ear and mastoid process) |
| **L00-L98** | Other natural | O | L00-L98 (Diseases of the skin and subcutaneous tissue) |
| **M00-M99** | Other natural | O | M00-M99 (Diseases of the musculoskeletal system and connective tissue) |
| **N00-N98** | Other natural | O | N00-N98 (Diseases of the genitourinary system) |
| **O00-O99** | Other natural | O | O00-O99 (Pregnancy, childbirth and the puerperium) |
| **P00-P96** | Other natural | O | P00-P96 (Certain conditions originating in the perinatal period) |
| **Q00-Q99** | Other natural | O | Q00-Q99 (Congenital malformations, deformations and chromosomal abnormalities) |
| **R00-R09** | Other natural | O | R00-R09 (Symptoms and signs involving the circulatory and respiratory systems) |
| **R10-R19** | Other natural | O | R10-R19 (Symptoms and signs involving the digestive system and abdomen) |
| **R20-R23** | Other natural | O | R20-R23 (Symptoms and signs involving the skin and subcutaneous tissue) |
| **R25-R29** | Other natural | O | R25-R29 (Symptoms and signs involving the nervous and musculoskeletal systems) |
| **R30-R39** | Other natural | O | R30-R39 (Symptoms and signs involving the urinary system) |
| **R40-R46** | Other natural | O | R40-R46 (Symptoms and signs involving cognition, perception, emotional state and behaviour) |
| **R47-R49** | Other natural | O | R47-R49 (Symptoms and signs involving speech and voice) |
| **R50-R68** | Other natural | O | R50-R68 (General symptoms and signs) |
| **R70** | Other natural | O | R70 (Elevated erythrocyte sedimentation rate and abnormality of plasma viscosity) |
| **R71** | Other natural | O | R71 (Abnormality of red blood cells) |
| **R72** | Other natural | O | R72 (Abnormality of white blood cells, not elsewhere classified) |
| **R73** | Other natural | O | R73 (Elevated blood glucose level) |
| **R74** | Other natural | O | R74 (Abnormal serum enzyme levels) |
| **R75** | Other natural | O | R75 (Laboratory evidence of human immunodeficiency virus [HIV]) |
| **R76** | Other natural | O | R76 (Other abnormal immunological findings in serum) |
| **R77** | Other natural | O | R77 (Other abnormalities of plasma proteins) |
| **R781** | Other natural | O | R78.1 (Finding of opiate drug in blood) |
| **R782** | Other natural | O | R78.2 (Finding of cocaine in blood) |
| **R783** | Other natural | O | R78.3 (Finding of hallucinogen in blood) |
| **R784** | Other natural | O | R78.4 (Finding of other drugs of addictive potential in blood) |
| **R785** | Other natural | O | R78.5 (Finding of psychotropic drug in blood) |
| **R786** | Other natural | O | R78.6 (Finding of steroid agent in blood) |
| **R787** | Other natural | O | R78.7 (Finding of abnormal level of heavy metals in blood) |
| **R788** | Other natural | O | R78.8 (Finding of other specified substances, not normally found in blood) |
| **R789** | Other natural | O | R78.9 (Finding of unspecified substance, not normally found in blood) |
| **R79** | Other natural | O | R79 (Other abnormal findings of blood chemistry) |
| **R80-R82** | Other natural | O | R80-R82 (Abnormal findings on examination of urine, without diagnosis) |
| **R83-R89** | Other natural | O | R83-R89 (Abnormal findings on examination of other body fluids, substances and tissues, without diagnosis) |
| **R90-R94** | Other natural | O | R90-R94 (Abnormal findings on diagnostic imaging and in function studies, without diagnosis) |
| **R95-R99** | Other natural | O | R95-R99 (Ill-defined and unknown causes of mortality) |
| **U04** | Other natural | O | U04 (Severe acute respiratory syndrome [SARS]) |
| **U070** | Other natural | O | U07.0 (Vaping related disorder) |
| **J00-J98** | Respiratory | R | J00-J98 (Diseases of the respiratory system) |
| **X66** | Suicide | S | X66 (Intentional self-poisoning by and exposure to organic solvents and halogenated hydrocarbons and their vapours) |
| **X67** | Suicide | S | X67 (Intentional self-poisoning by and exposure to other gases and vapours) |
| **X68** | Suicide | S | X68 (Intentional self-poisoning by and exposure to pesticides) |
| **X69** | Suicide | S | X69 (Intentional self-poisoning by and exposure to other and unspecified chemicals and noxious substances) |
| **X70** | Suicide | S | X70 (Intentional self-harm by hanging, strangulation and suffocation) |
| **X71** | Suicide | S | X71 (Intentional self-harm by drowning and submersion) |
| **X72** | Suicide | S | X72 (Intentional self-harm by handgun discharge) |
| **X73** | Suicide | S | X73 (Intentional self-harm by rifle, shotgun and larger firearm discharge) |
| **X74** | Suicide | S | X74 (Intentional self-harm by other and unspecified firearm discharge) |
| **X75** | Suicide | S | X75 (Intentional self-harm by explosive material) |
| **X76** | Suicide | S | X76 (Intentional self-harm by smoke, fire and flames) |
| **X77** | Suicide | S | X77 (Intentional self-harm by steam, hot vapours and hot objects) |
| **X78** | Suicide | S | X78 (Intentional self-harm by sharp object) |
| **X79** | Suicide | S | X79 (Intentional self-harm by blunt object) |
| **X80** | Suicide | S | X80 (Intentional self-harm by jumping from a high place) |
| **X81** | Suicide | S | X81 (Intentional self-harm by jumping or lying before moving object) |
| **X82** | Suicide | S | X82 (Intentional self-harm by crashing of motor vehicle) |
| **X83** | Suicide | S | X83 (Intentional self-harm by other specified means) |
| **X84** | Suicide | S | X84 (Intentional self-harm by unspecified means) |
| **Y870** | Suicide | S | Y87.0 (Sequelae of intentional self-harm) |
| **U01** | Other external | O | U01 (Terrorism - Assault (homicide) ) |
| **U02** | Other external | O | U02 (Sequelae of terrorism) |
| **U03** | Other external | O | U03 (Terrorism Intentional (Suicide)) |
| **W00-W19** | Falls | F | W00-W19 (Falls) |
| **W20-W49** | Other external | O | W20-W49 (Exposure to inanimate mechanical forces) |
| **W50-W64** | Other external | O | W50-W64 (Exposure to animate mechanical forces) |
| **W65-W74** | Other external | O | W65-W74 (Accidental drowning and submersion) |
| **W75-W84** | Other external | O | W75-W84 (Other accidental threats to breathing) |
| **W85-W99** | Other external | O | W85-W99 (Exposure to electric current, radiation and extreme ambient air temperature and pressure) |
| **X00-X09** | Other external | O | X00-X09 (Exposure to smoke, fire and flames) |
| **X10-X19** | Other external | O | X10-X19 (Contact with heat and hot substances) |
| **X20-X29** | Other external | O | X20-X29 (Contact with venomous animals and plants) |
| **X30-X39** | Other external | O | X30-X39 (Exposure to forces of nature) |
| **X46** | Other external | O | X46 (Accidental poisoning by and exposure to organic solvents and halogenated hydrocarbons and their vapours) |
| **X47** | Other external | O | X47 (Accidental poisoning by and exposure to other gases and vapours) |
| **X48** | Other external | O | X48 (Accidental poisoning by and exposure to pesticides) |
| **X49** | Other external | O | X49 (Accidental poisoning by and exposure to other and unspecified chemicals and noxious substances) |
| **X50-X57** | Other external | O | X50-X57 (Overexertion, travel and privation) |
| **Y16** | Other external | O | Y16 (Poisoning by and exposure to organic solvents and halogenated hydrocarbons and their vapours, undetermined intent) |
| **Y17** | Other external | O | Y17 (Poisoning by and exposure to other gases and vapours, undetermined intent) |
| **Y18** | Other external | O | Y18 (Poisoning by and exposure to pesticides, undetermined intent) |
| **Y19** | Other external | O | Y19 (Poisoning by and exposure to other and unspecified chemicals and noxious substances, undetermined intent) |
| **Y20** | Other external | O | Y20 (Hanging, strangulation and suffocation, undetermined intent) |
| **Y21** | Other external | O | Y21 (Drowning and submersion, undetermined intent) |
| **Y22** | Other external | O | Y22 (Handgun discharge, undetermined intent) |
| **Y23** | Other external | O | Y23 (Rifle, shotgun and larger firearm discharge, undetermined intent) |
| **Y24** | Other external | O | Y24 (Other and unspecified firearm discharge, undetermined intent) |
| **Y25** | Other external | O | Y25 (Contact with explosive material, undetermined intent) |
| **Y26** | Other external | O | Y26 (Exposure to smoke, fire and flames, undetermined intent) |
| **Y27** | Other external | O | Y27 (Contact with steam, hot vapours and hot objects, undetermined intent) |
| **Y28** | Other external | O | Y28 (Contact with sharp object, undetermined intent) |
| **Y29** | Other external | O | Y29 (Contact with blunt object, undetermined intent) |
| **Y30** | Other external | O | Y30 (Falling, jumping or pushed from a high place, undetermined intent) |
| **Y31** | Other external | O | Y31 (Falling, lying or running before or into moving object, undetermined intent) |
| **Y32** | Other external | O | Y32 (Crashing of motor vehicle, undetermined intent) |
| **Y33** | Other external | O | Y33 (Other specified events, undetermined intent) |
| **Y34** | Other external | O | Y34 (Unspecified event, undetermined intent) |
| **Y86** | Other external | O | Y86 (Sequelae of other accidents) |
| **Y872** | Other external | O | Y87.2 (Sequelae of events of undetermined intent) |
| **Y88** | Other external | O | Y88 (Sequelae with surgical and medical care as external cause) |
| **Y89** | Other external | O | Y89 (Sequelae of other external causes) |
| **V01-V99** | Transport | T | V01-V99 (Transport accidents) |
| **Y85** | Transport | T | Y85 (Sequelae of transport accidents) |
| **T36-T50** | Drug poisoning | T | T36-T50 (Poisoning by drugs and biological substances) |
| **S00-S99** | Other external | S | S00-S99 (Injuries to specific body regions) |
| **T00-T35** | Other external | T | T00-T35 (Injuries involving multiple/unspecified body regions and burns) |
| **T51-T65** | Other external | T | T51-T65 (Toxic effects of substances chiefly nonmedicinal as to source) |
| **T66-T78** | Other external | T | T66-T78 (Other and unspecified effects of external causes) |
| **T79** | Other external | T | T79 (Certain early complications of trauma) |
| **T80-T89** | Other external | T | T80-T89 (Complications of surgical and medical care NEC) |
| **T90-T99** | Other external | T | T90-T99 (Sequelae of injuries and other external causes) |
| **V00** | Transport | V | V00 (Pedestrian conveyance accident) |
| **X58** | Other external | X | X58 (Exposure to other specified factors) |
| **X59** | Other external | X | X59 (Exposure to unspecified factor) |
| **Y35-Y39** | Other external | Y | Y35-Y39 (Legal intervention and operations of war) |
| **Y40-Y59** | Other external | Y | Y40-Y59 (Drugs and biological substances causing adverse effects in therapeutic use) |
| **Y60-Y69** | Other external | Y | Y60-Y69 (Misadventures to patients during surgical and medical care) |
| **Y70-Y82** | Other external | Y | Y70-Y82 (Medical devices associated with adverse incidents) |
| **Y83-Y84** | Other external | Y | Y83-Y84 (Surgical and other medical procedures as cause of abnormal reaction) |
| **Y90-Y99** | Other external | Y | Y90-Y99 (Supplementary factors related to causes of morbidity and mortality) |

### Supplementary Figures

**Supplementary Figure S1.**
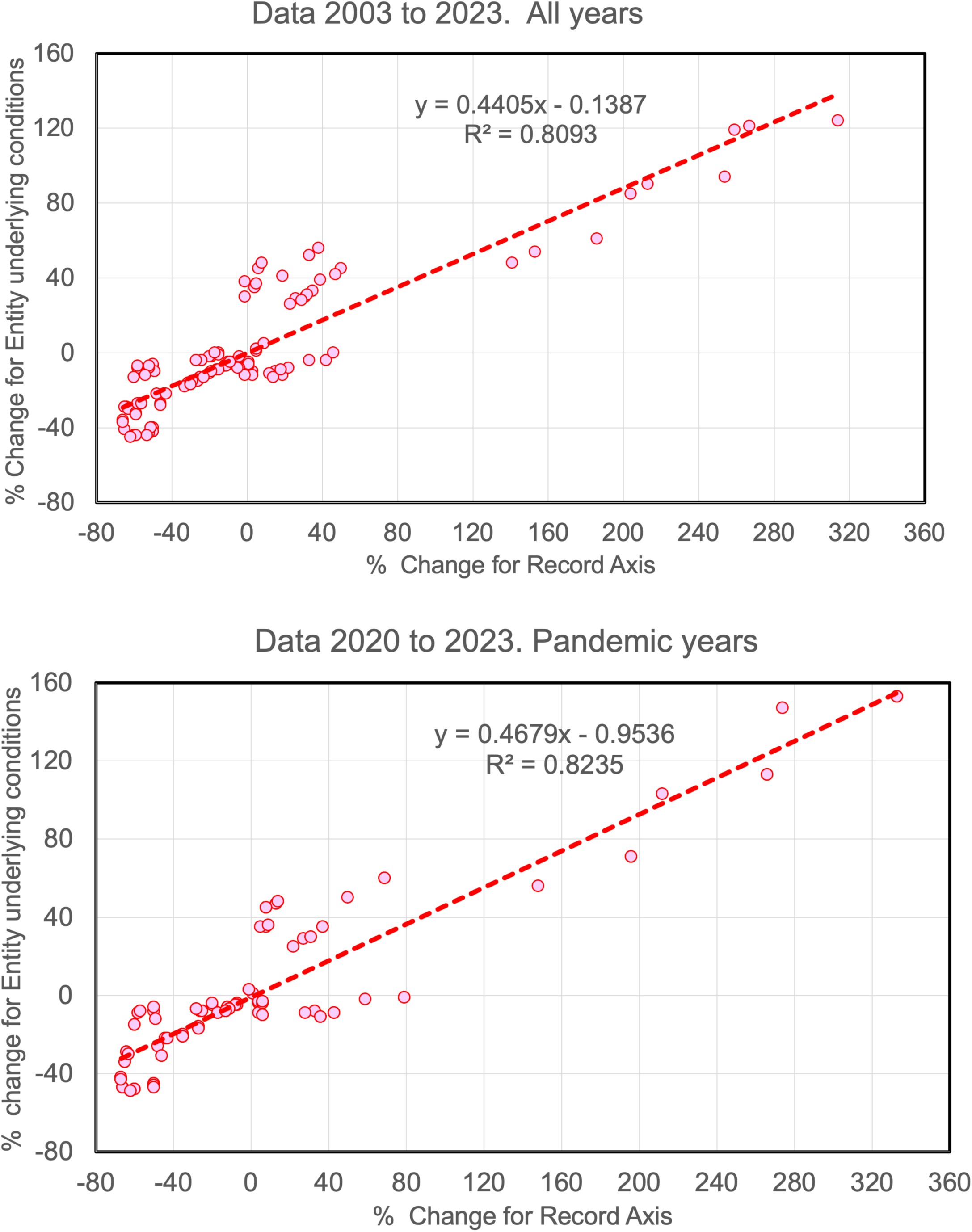
Showing how the percent changes of the broad disease category counts depend on the source of data. Along the x-axis we show data from the Record Axis. Along the y-axis we show data that combines the underlying cause-of-death from the Record Axis with the contributing causes from Part II of the Entity Axis.

**Supplementary Figure S2.**
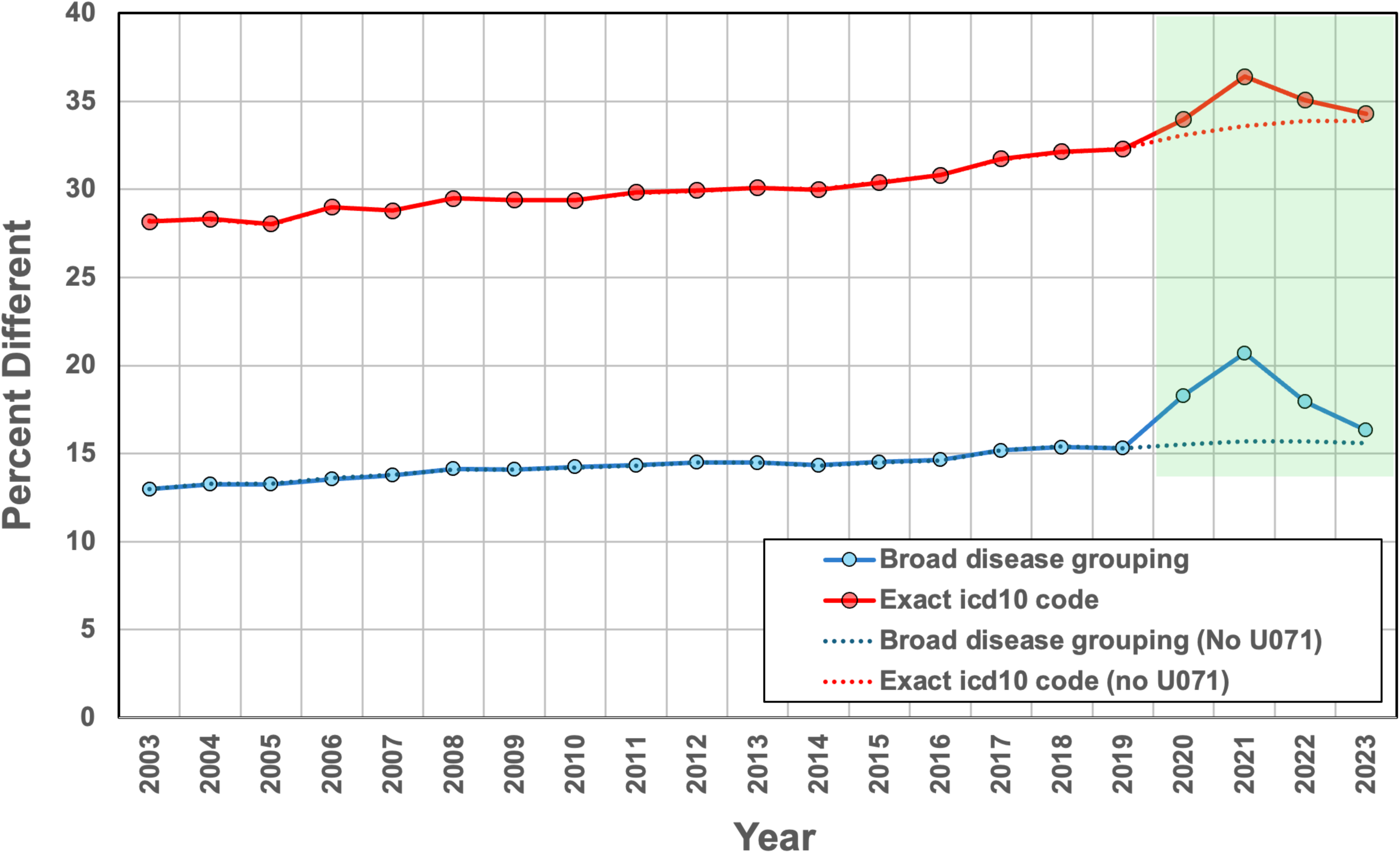
Increase in cause-of-death coding differences between death certifier-assigned and NCHS/ACME-assigned underlying cause of death over time and especially during the early years of the COVID-19 pandemic. Data shown for ICD-10 coding and broad disease categories with and excluding the key COVID-19 code. This data is shown in Table S5B.

**Supplementary Figure S3.**
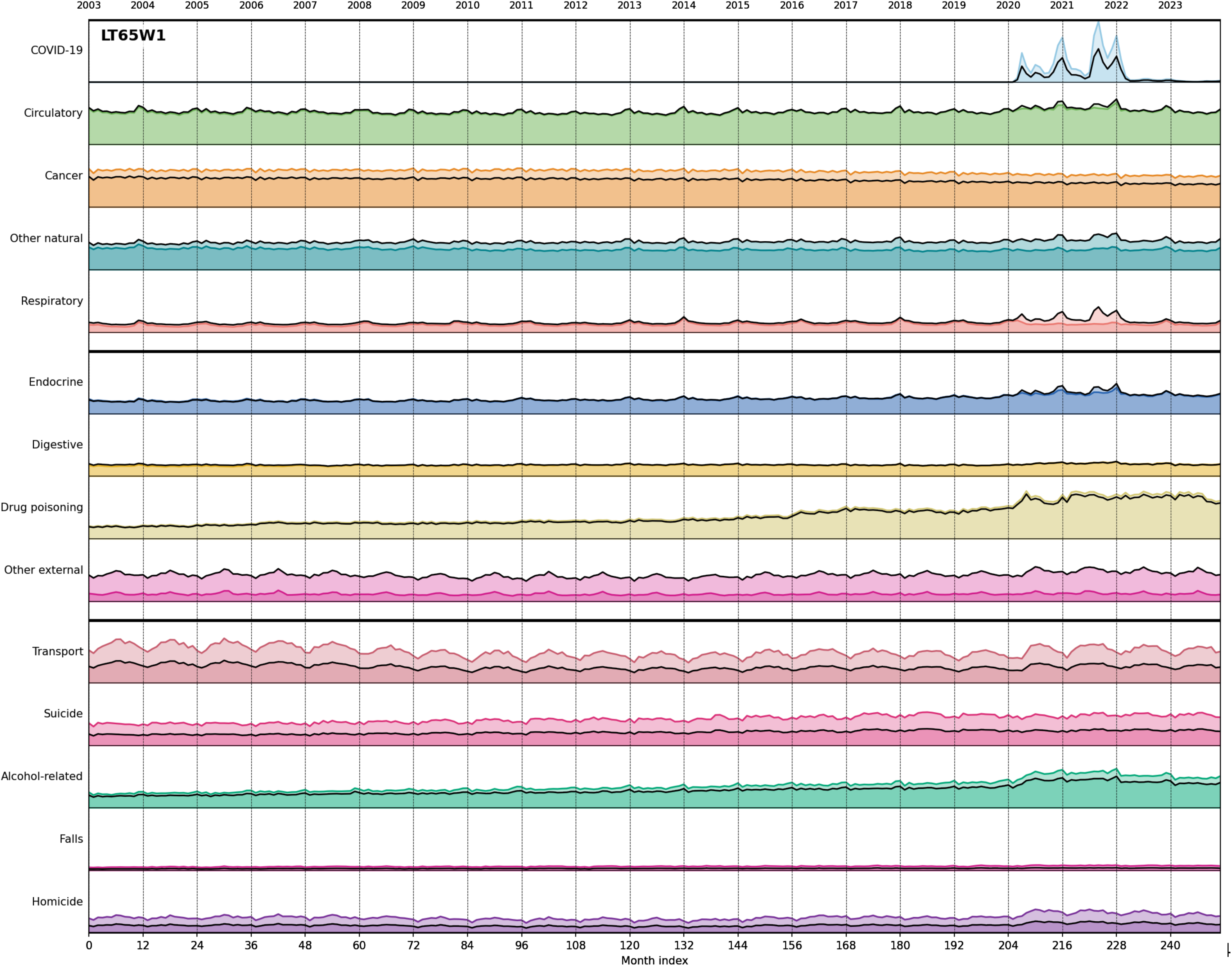
**(LT65W1).** Population-normalized monthly deaths across 14 broad categories during 2003-2023 for non-elderly people (less than 65 years old). The panels are in different scale. Light line: unweighted W0. Continuous thick line: W1 weights. Disease categories are grouped by fixed Y-axis scales to allow visual comparison across panels. The top band (COVID-19, Circulatory, Cancer, Other natural, Respiratory) uses 0-26,000 deaths/month; the middle band (Endocrine, Digestive, Drug poisoning, Other external) uses 0-12,000 deaths/month; the bottom band (Transport, Suicide, Alcohol-related, Falls, Homicide) uses 0-6,000 deaths/month. All values are population-scaled to 2023 levels.

**Supplementary Figure S4.**
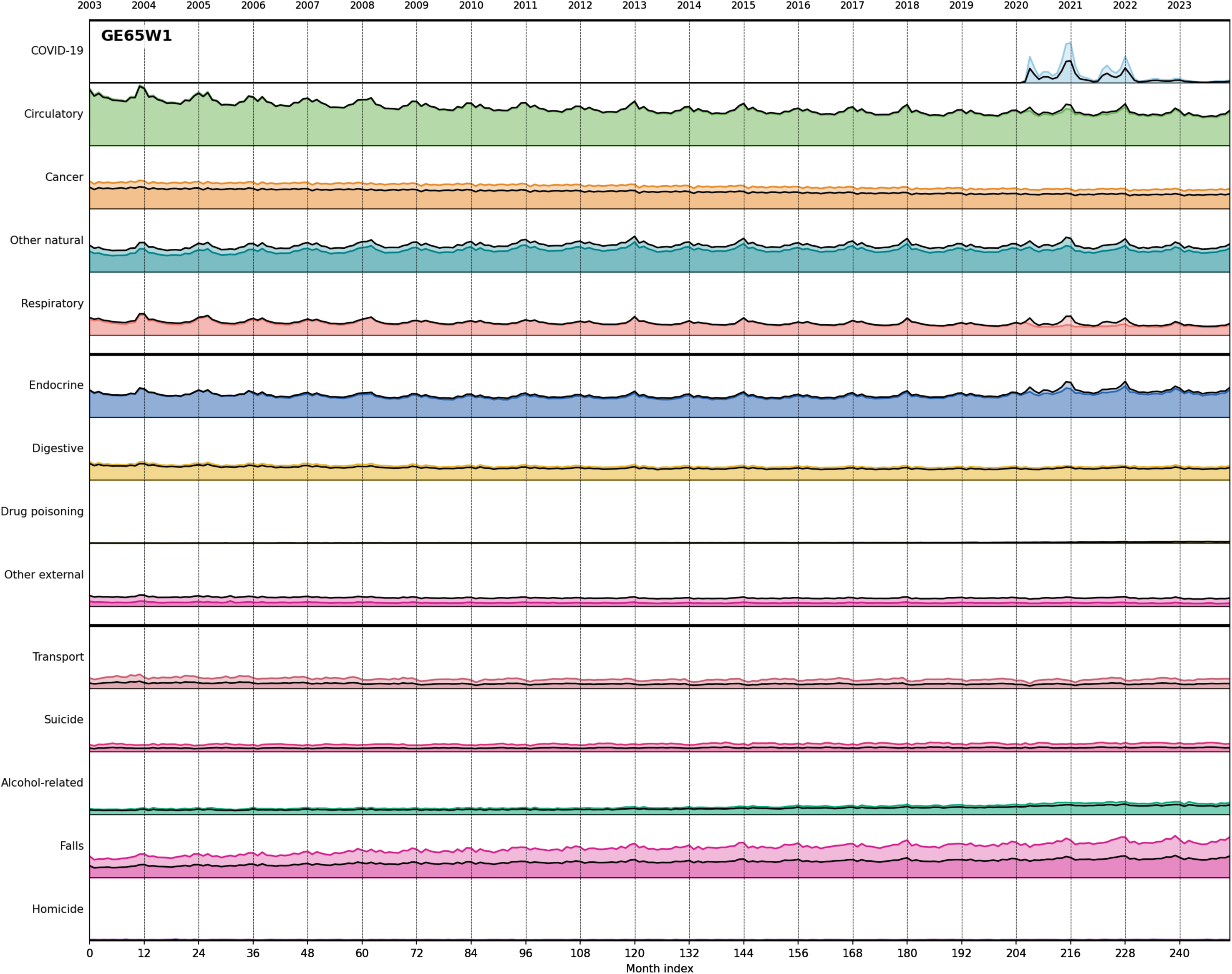
**(GE65W1).** Population-normalized monthly deaths across 14 broad categories during 2003-2023 for elderly people (age 65 years and over). The panels are in different scale. Light line: unweighted W0. Continuous thick line: W1 weights. Disease categories are grouped by fixed Y-axis scales to allow visual comparison across panels. The top band (COVID-19, Circulatory, Cancer, Other natural, Respiratory) uses 0-130,000 deaths/month; the middle band (Endocrine, Digestive, Drug poisoning, Other external) uses 0-25,000 deaths/month; the bottom band (Transport, Suicide, Alcohol-related, Falls, Homicide) uses 0-6,000 deaths/month.

**Supplementary Figure S5.**
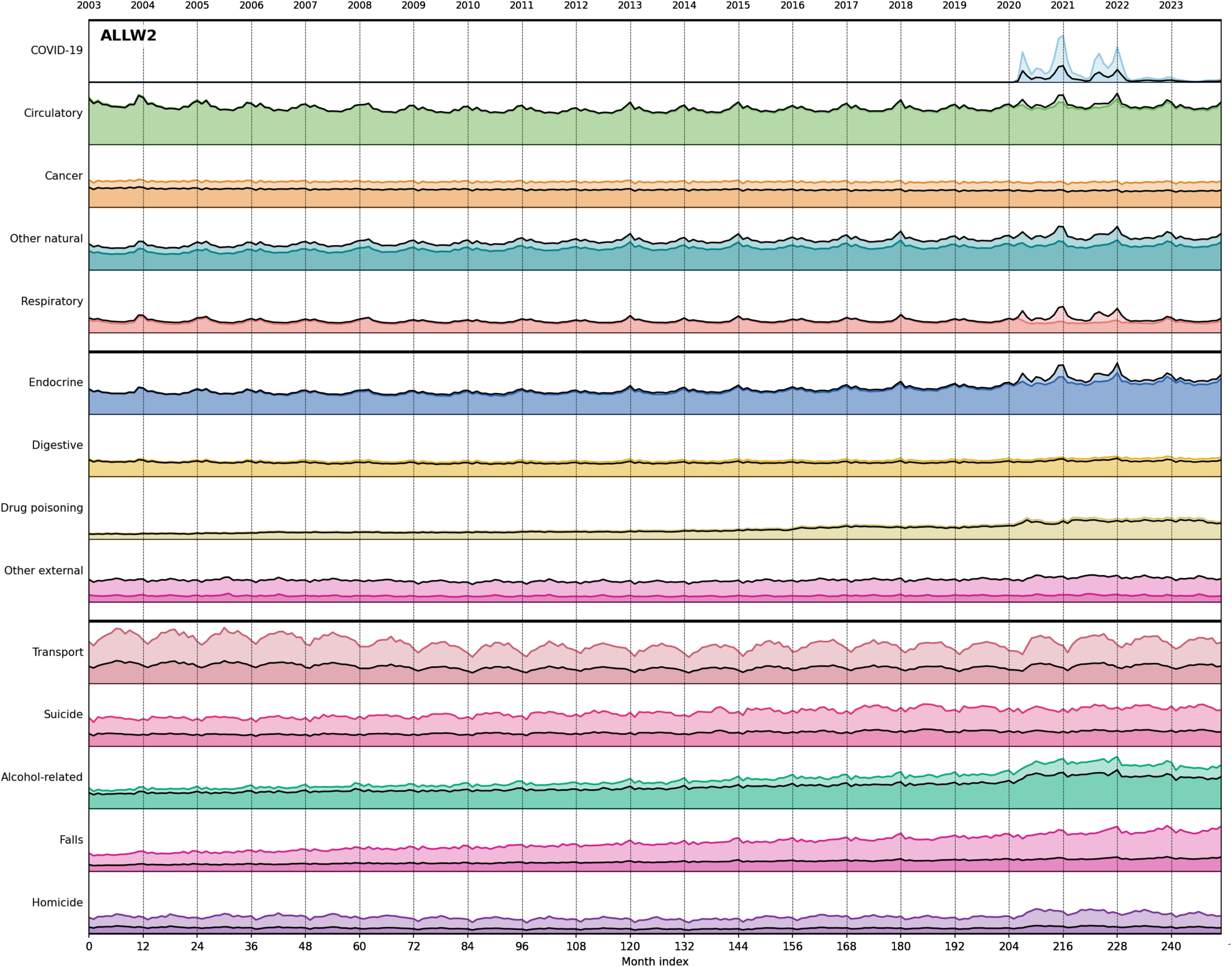
**(ALLW2).** Population-normalized monthly deaths across 14 broad categories during 2003-2023. The panels are in different scale. Light line: unweighted W0. Continuous thick line: W2 weights. Disease categories are grouped by fixed Y-axis scales to allow visual comparison across panels. The top band (COVID-19, Circulatory, Cancer, Other natural, Respiratory) uses 0-130,000 deaths/month; the middle band (Endocrine, Digestive, Drug poisoning, Other external) uses 0-25,000 deaths/month; the bottom band (Transport, Suicide, Alcohol-related, Falls, Homicide) uses 0-6,000 deaths/month. All values are population-scaled to 2023 levels.

**Supplementary Figure S6.**
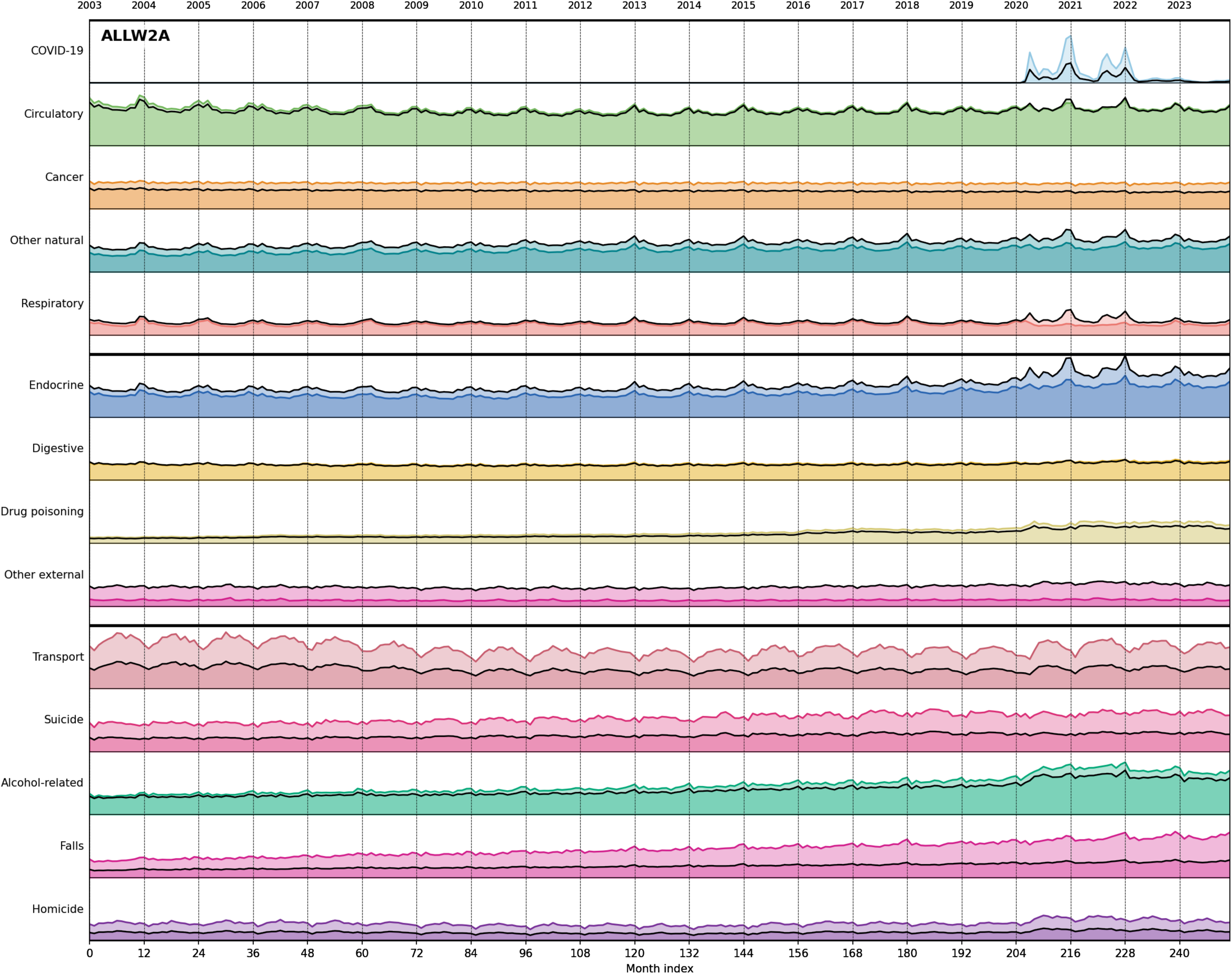
**(ALLW2A).** Population-normalized monthly deaths across 14 broad categories during 2003-2023. The panels are in different scale. Light line: unweighted W0. Continuous thick line: W2A weights. Disease categories are grouped by fixed Y-axis scales to allow visual comparison across panels. The top band (COVID-19, Circulatory, Cancer, Other natural, Respiratory) uses 0-130,000 deaths/month; the middle band (Endocrine, Digestive, Drug poisoning, Other external) uses 0-25,000 deaths/month; the bottom band (Transport, Suicide, Alcohol-related, Falls, Homicide) uses 0-6,000 deaths/month. All values are population-scaled to 2023 levels.

**Supplementary Figure S7.**
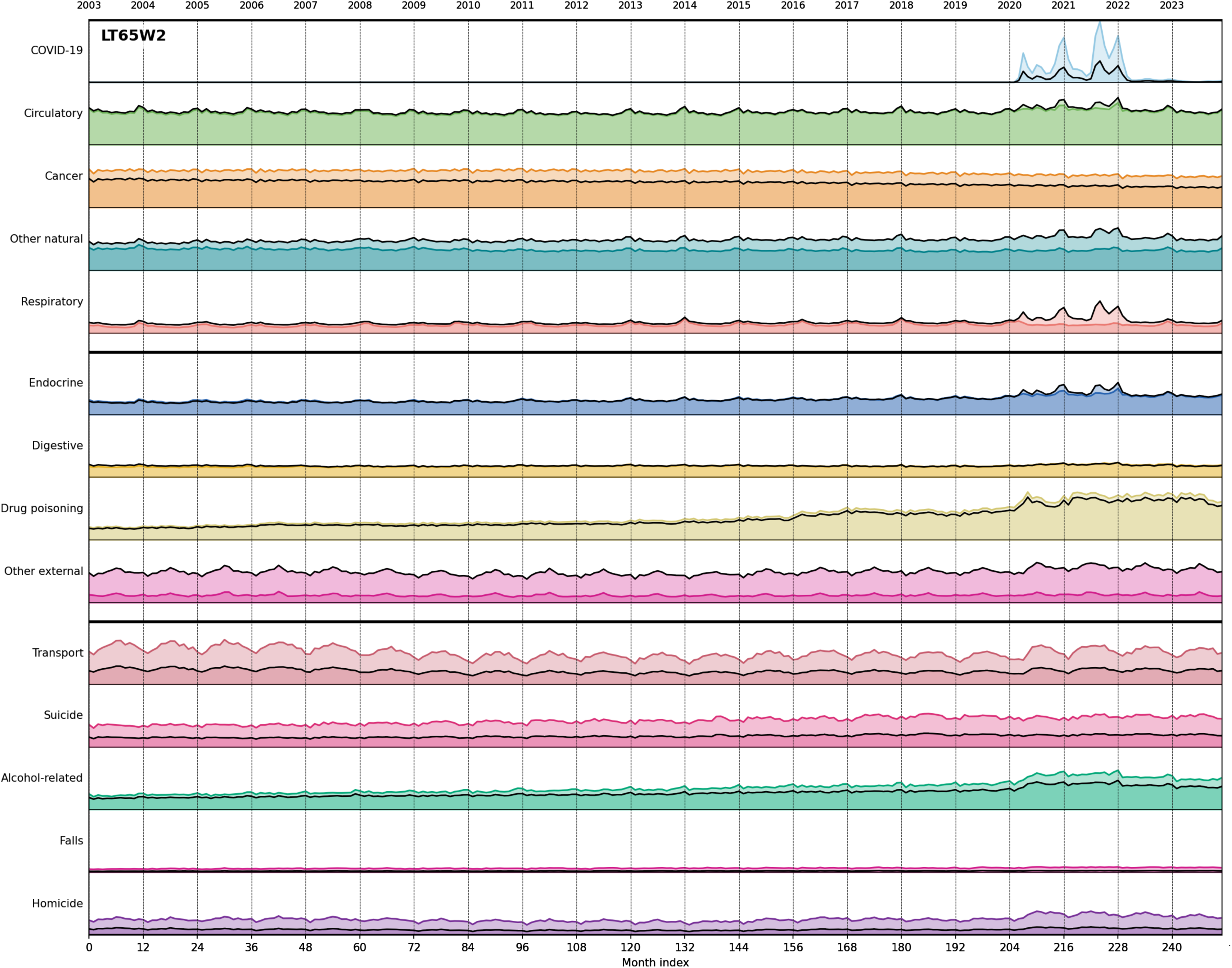
**(LT65W2).** Population-normalized monthly deaths across 14 broad categories during 2003-2023 for non-elderly people (less than 65 years old). The panels are in different scale. Light line: unweighted W0. Continuous thick line: W2 weights. Disease categories are grouped by fixed Y-axis scales to allow visual comparison across panels. The top band (COVID-19, Circulatory, Cancer, Other natural, Respiratory) uses 0-26,000 deaths/month; the middle band (Endocrine, Digestive, Drug poisoning, Other external) uses 0-12,000 deaths/month; the bottom band (Transport, Suicide, Alcohol-related, Falls, Homicide) uses 0-6,000 deaths/month. All values are population-scaled to 2023 levels.

**Supplementary Figure S8.**
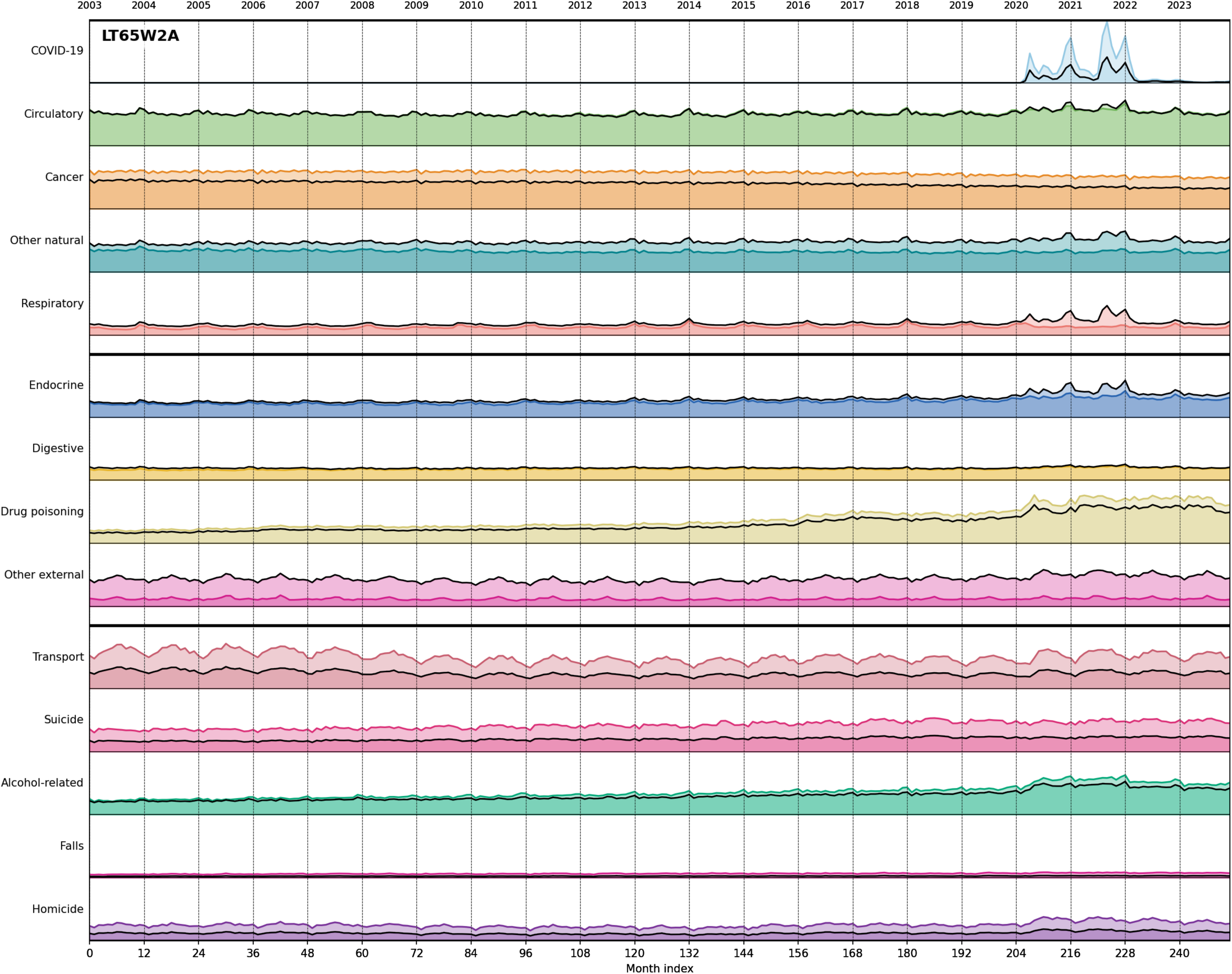
**(LT65W2A).** Population-normalized monthly deaths across 14 broad categories during 2003-2023 for non-elderly people (less than 65 years old). The panels are in different scale. Light line: unweighted W0. Continuous thick line: W2A weights. Disease categories are grouped by fixed Y-axis scales to allow visual comparison across panels. The top band (COVID-19, Circulatory, Cancer, Other natural, Respiratory) uses 0-26,000 deaths/month; the middle band (Endocrine, Digestive, Drug poisoning, Other external) uses 0-12,000 deaths/month; the bottom band (Transport, Suicide, Alcohol-related, Falls, Homicide) uses 0-6,000 deaths/month. All values are population-scaled to 2023 levels.

**Supplementary Figure S9.**
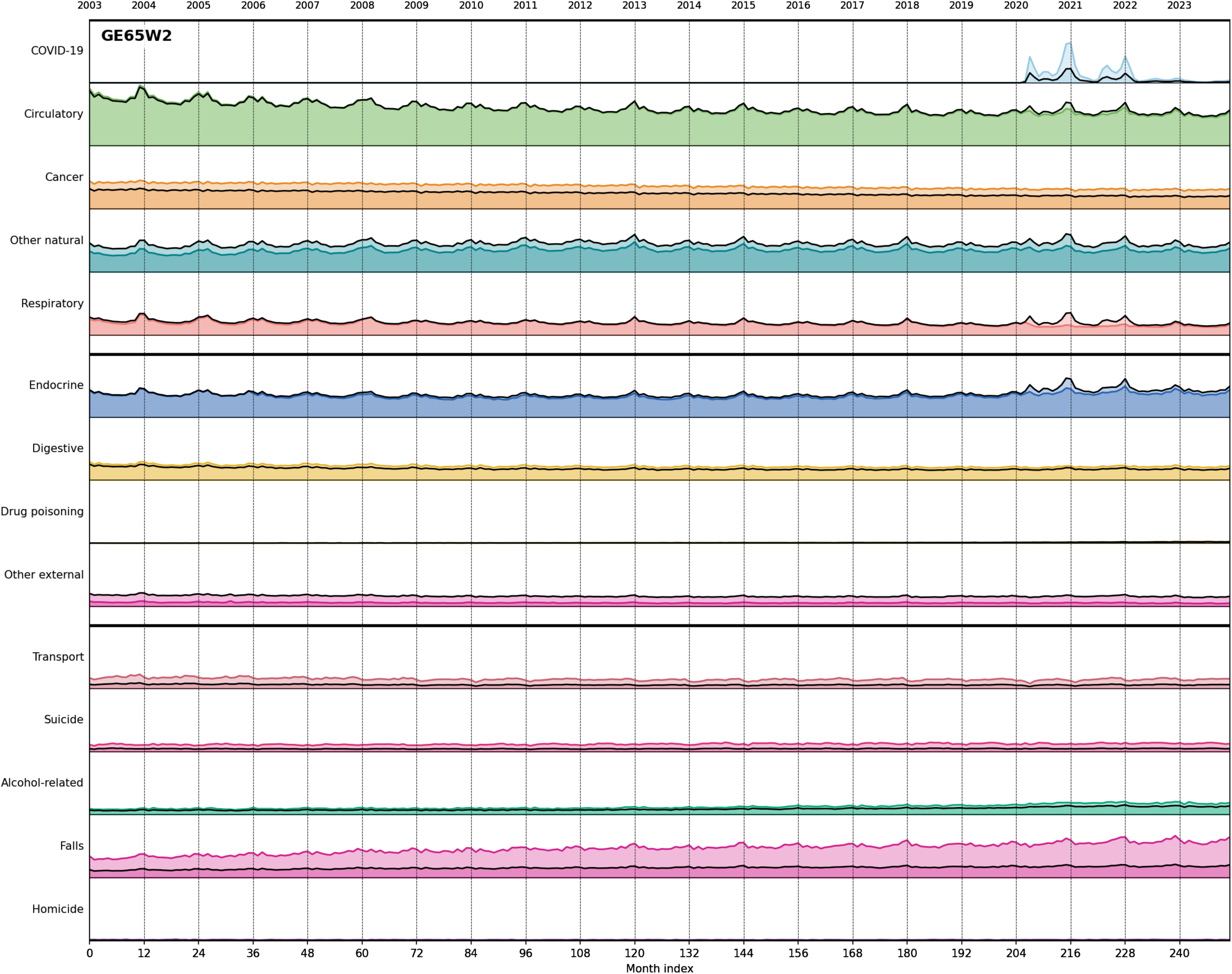
**(GE65W2).** Population-normalized monthly deaths across 14 broad categories during 2003-2023 for elderly people (age 65 years and over). The panels are in different scale. Light line: unweighted W0. Continuous thick line: W2 weights. Disease categories are grouped by fixed Y-axis scales to allow visual comparison across panels. The top band (COVID-19, Circulatory, Cancer, Other natural, Respiratory) uses 0-130,000 deaths/month; the middle band (Endocrine, Digestive, Drug poisoning, Other external) uses 0-25,000 deaths/month; the bottom band (Transport, Suicide, Alcohol-related, Falls, Homicide) uses 0-6,000 deaths/month. All values are population-scaled to 2023 levels.

**Supplementary Figure S10.**
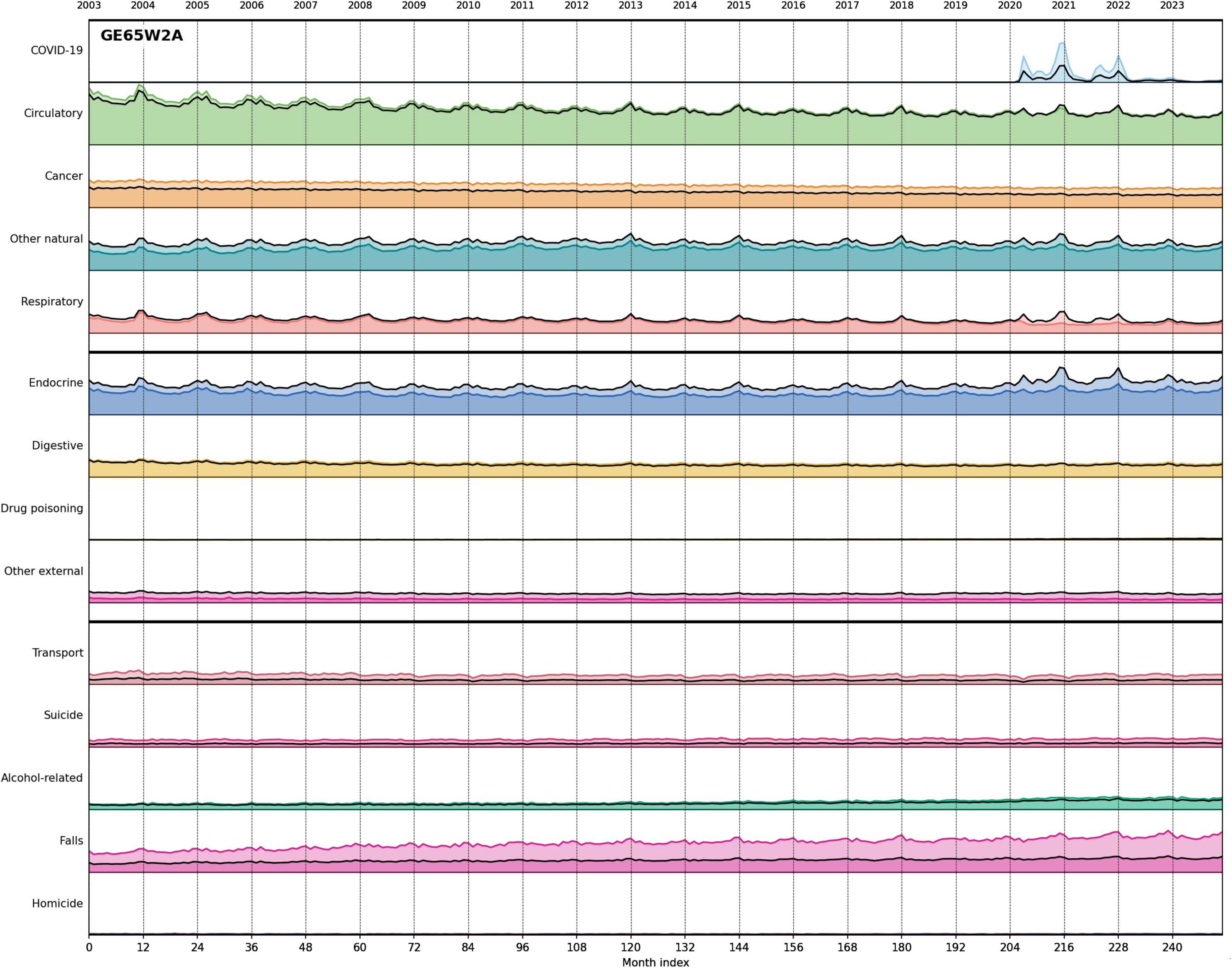
**(GE65W2A).** Population-normalized monthly deaths across 14 broad categories during 2003-2023 for elderly people (age 65 years and over). The panels are in different scale. Light line: unweighted W0. Continuous thick line: W2A weights. Disease categories are grouped by fixed Y-axis scales to allow visual comparison across panels. The top band (COVID-19, Circulatory, Cancer, Other natural, Respiratory) uses 0-130,000 deaths/month; the middle band (Endocrine, Digestive, Drug poisoning, Other external) uses 0-25,000 deaths/month; the bottom band (Transport, Suicide, Alcohol-related, Falls, Homicide) uses 0-6,000 deaths/month. All values are population-scaled to 2023 levels.

**Supplementary Figure S11.**
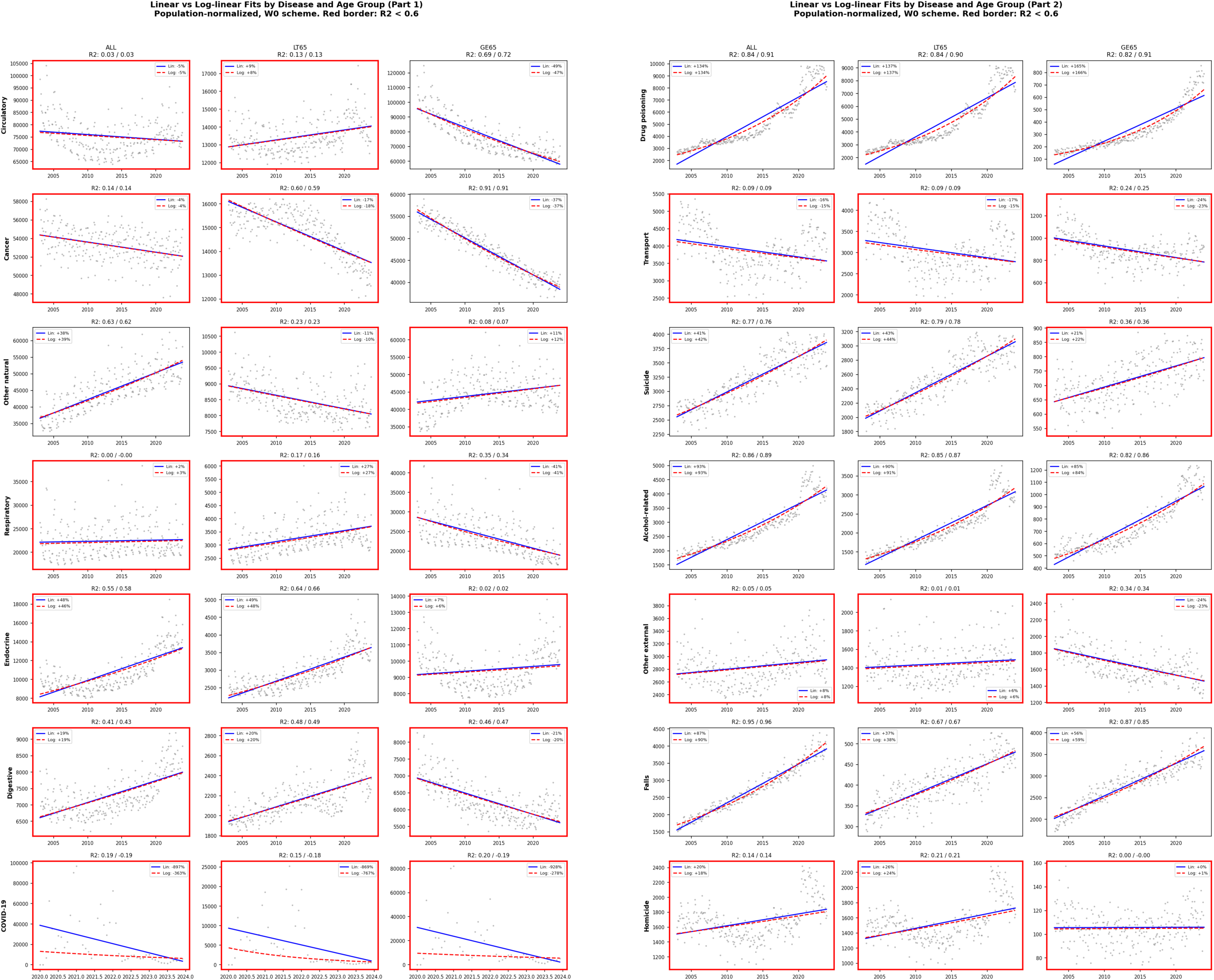
Linear and logarithmic fits to the population-normalized monthly deaths for the three age buckets (ALL, LT65 for people under 65 and GE65 for elderly people aged 65 years and over) across 14 broad categories during 2003-2023. Plots with unacceptable fits (R2 < 0.6, CC <. 0.77) have red plot boundaries. They are the majority occurring in 26 out of 42 cases. The other 16 plots show clear linear change over 21 years, with the possible exception of Drug poisoning, which grows somewhat exponentially.

